# Epigenetic Reprogramming Mediates Monocyte and Heterologous T Cell-derived Cytokine Responses after BCG Vaccination

**DOI:** 10.1101/2024.03.27.24304976

**Authors:** Cancan Qi, Zhaoli Liu, Gizem Kilic, Andrei S. Sarlea, Priya A. Debisarun, Xuan Liu, Yonatan Ayalew Mekonnen, Wenchao Li, Martin Grasshoff, Ahmed Alaswad, Apostolos Petkoglou, Valerie A.C.M. Koeken, Simone J.C.F.M. Moorlag, L. Charlotte J. de Bree, Vera P. Mourits, Leo A.B. Joosten, Yang Li, Mihai G. Netea, Cheng-Jian Xu

## Abstract

Epigenetic reprogramming plays an important role in shaping immune memory traits within both innate (trained immunity) and adaptive immune cells following Bacillus Calmette-Guérin (BCG) vaccination. However, the precise impact of dynamic DNA methylation alterations on immunological responses after BCG vaccination remains inadequately elucidated. To address this knowledge gap, we conducted a comprehensive study by integrating longitudinal analysis and systems biology approaches. We established a cohort of 284 healthy Dutch individuals, capturing data on genetics, cytokine responses to *ex vivo* stimulation and genome-wide DNA methylation at baseline, as well as at 14 days and 90 days after BCG vaccination. Our findings revealed distinct patterns of DNA methylation alternations in the short- and long-term following BCG vaccination. Moreover, we established that baseline DNA methylation profiles exert influence on the change in interferon-γ (IFN-γ) production upon heterologous (*Staphylococcus aureus)* stimulation before and after BCG vaccination. Specifically, we identified the regulation of kisspeptin as a novel pathway implicated in the modulation of IFN-γ production, and this finding has been substantiated through experiment validation. We also observed associations between BCG-induced DNA methylation changes and increased IFN-γ and Interleukin-1 β (IL-1β) production upon *S. aureus* stimulation. Interestingly, by integrating with genetic, epigenetic, and cytokine response data from the same individuals, mediation analysis demonstrated that most of the identified DNA methylation changes played a mediating role between genetic variants and cytokine responses, for example, the changes of cg21375332 near *SLC12A3* gene mediated the regulation of genetic variants on IFN-γ changes after BCG vaccination. Sex-specific effects consistently manifested in DNA methylation changes after BCG vaccination and in the association between baseline methylation and cytokine responses. Together, our findings provide deeper insights into immune response mechanisms, crucial for developing effective epigenetic-based medical interventions for personalized medicine.

## Introduction

Host immune responses are traditionally classified into innate and adaptive, with only the latter initially thought to have the ability to develop immunological memory. However, in recent years, a growing body of evidence has shown that innate immunity can also exhibit memory characteristics^1^. Studies have shown explicitly that Bacillus Calmette-Guérin (BCG) vaccination can induce innate immune memory through epigenetic reprogramming of myeloid cells and their bone marrow progenitors ^2–6^. This non-specific innate immune memory can provide cross-protection against unrelated-pathogen and is referred to as “trained immunity” ^7,8^. In addition, BCG vaccination induces classical adaptive immune memory, with T-cell-derived IFN-γ responses being relevant for protection against tuberculosis ^2^.

A distinguishing feature of the trained innate immune cells is their ability to mount a stronger transcriptional response compared to naïve cells when challenged with a pathogen. Recent research has identified various factors that can influence the induction of trained immunity, such as circulating inflammatory proteins ^9^, metabolites^10^ and the transcriptomic profiles of immune cells ^11–13^, and the gut microbiome ^14^. Studies have shown that after being vaccinated with BCG, systematic inflammation is reduced at both the protein and transcription levels ^9,11,12,15^. Additionally, the production of cytokine following BCG vaccination is associated with the abundance of microbial genomes, which in turn influences circulating metabolites ^14^.

One of the key molecular mechanisms involved in the induction of trained immunity is the epigenetic reprogramming of immune cells. Chromatin accessibility and histone marks have been the mechanisms most studied in this regard ^7,16^. Another important mechanism for epigenetic gene transcription regulation is through modulation of DNA methylation. Previous studies have demonstrated the association of DNA methylation with infection and immune memory ^17,18^, showing how infection-induced changes in DNA methylation regulate the transcriptional response to infection and contribute to short-term memory in innate immune cells ^17^. Demethylation of enhancer elements mediated by transcription factors (TFs) binding may allow for a faster response to a secondary infection and thus plays a crucial role in innate immune memory ^19^. More recently, studies have shown that BCG vaccination leads to changes in DNA methylation ^20^ in monocytes and T cells of children ^21^. We have previously suggested that DNA methylation changes after BCG vaccination in adults as well ^22^, but a comprehensive analysis of the impact of DNA methylation on innate immune memory is missing. To better understand the molecular basis of trained immunity, we investigated whether DNA methylation plays an important role in this process.

In this study, we hypothesize that *in vivo* BCG vaccination induces changes in DNA methylation associated with a trained immunity response. To test this hypothesis, we longitudinally assessed *ex vivo* cytokine production upon *Staphylococcus aureus* stimulation, and genome-wide DNA methylation at baseline, 14 days, and 90 days after BCG vaccination in a cohort of 303 healthy volunteers from the Human Functional Genomics Project (300BCG cohort, www.humanfunctionalgenomics.org). We assessed the dynamic changes in DNA methylation following BCG vaccination and their association with cytokine responses using integrative system biological approach. We further validated our findings in an independent cohort (BCG booster trial ^23^) and a number of the identified pathways in *in-vitro* experiments.

## Results

### Study design and global DNA methylation variability

This study was conducted in the 300BCG cohort, and the details of the cohort have been previously reported^9^. In brief, 303 healthy volunteers received BCG vaccination and whole blood samples were collected before (T0), 14 days (T14), and 3 months (T90) after vaccination. The genome-wide DNA methylation profiles were then measured on high-quality DNA isolated from the whole blood using Illumina Epic arrays. Additionally, the production of cytokines (IL-1β, IL-6, TNF-α, IFN-γ) from peripheral blood mononuclear cells (PBMCs) was measured after stimulation with *Staphylococcus aureus* at T0 and T90. The fold change (FC) of cytokine production between T90 and T0, referred to as trained immunity markers in this study, was used as a measure of BCG-induced trained immune response. Plasma inflammatory protein concentrations were also obtained at three time points (Figure 1A). After rigorous quality control, downstream analyses included 284 subjects, consisting of 126 males and 158 females. The mean age was 25 years (ranging from 18 to 71), with a mean BMI of 22.48 kg/m^2^ (Table S1). Epigenome-wide association studies (EWAS) were performed on 751,564 high-quality probes from the three different time points of these 284 subjects. The EWAS aimed to examine: 1) the epigenetic modifications induced by BCG vaccination, and 2) the association between DNA methylation and changes of *ex vivo* cytokine production capacity induced by vaccination (Figure 1B).

**Figure 1.**
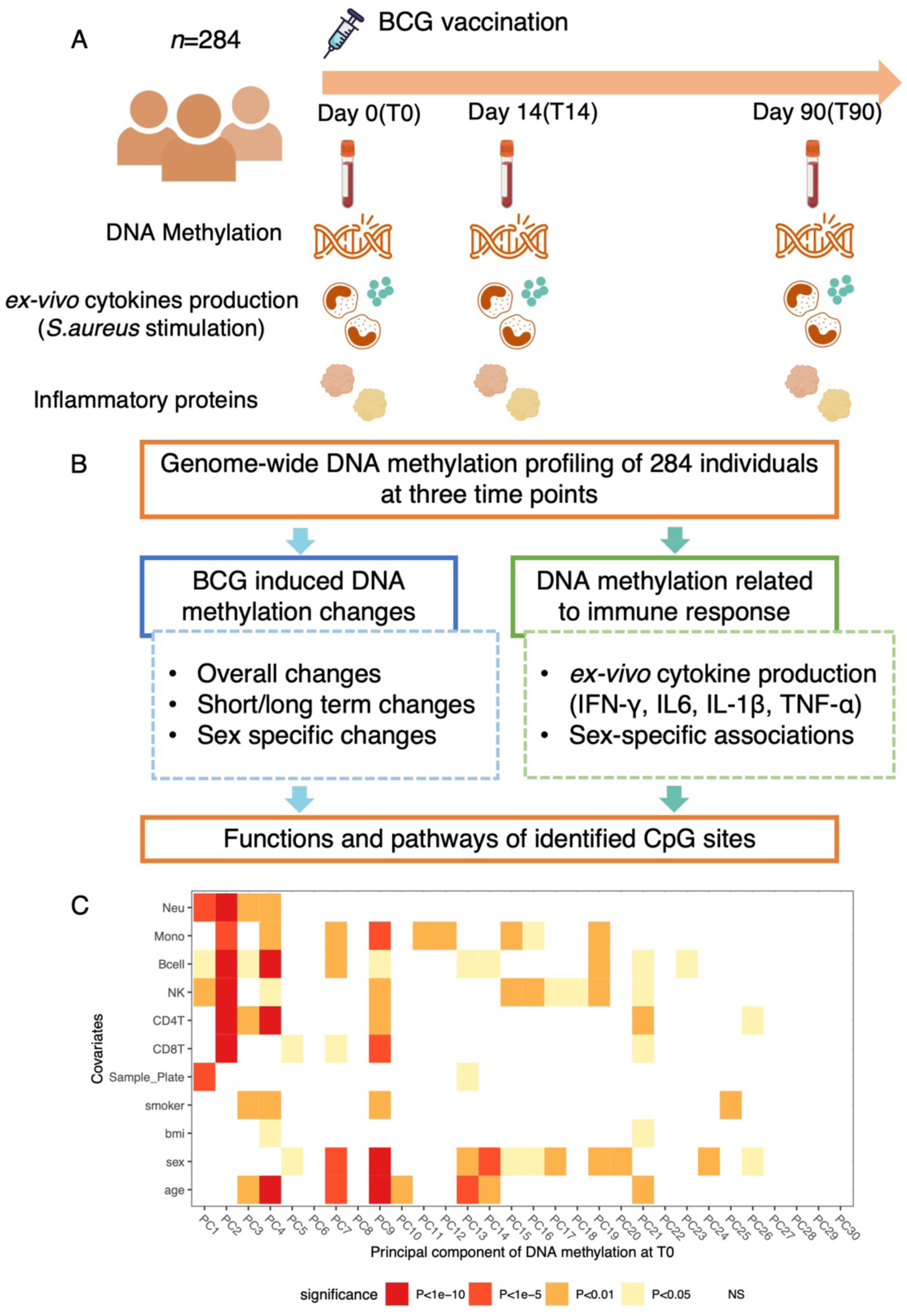
Study Design and data exploration. (A) Overview of the study design. (B) Main analysis in this study. (C) Heatmap of association between covariates, estimated cell proportions and DNA methylation at T0, which are represented as the top 30 PCs, capturing 35% of the variance. The association was performed with univariable linear regression model, different colors in the figure indicate different level of significance. BCG, Bacillus Calmette–Guérin; PC, principal component.

The univariate associations between the principal components (PCs) calculated from the whole blood DNA methylation data at each time point, covariates and estimated cell proportions^24^ are depicted in Figure 1C and S1. The first 30 PCs accounted for 35% of the total variation in the blood methylome at T0. The strongest associations were observed between the cell proportions, particularly neutrophils, and the top two DNA methylation PCs. The first DNA methylation PC was also associated with batch effects (sample plate), while age, sex, smoking behavior, and body mass index (BMI) showed no significant correlations with the top two PCs.

### BCG vaccination induced changes in DNA methylation

The changes in DNA methylation over time after BCG vaccination were assessed using a mixed effects model that accounted for age, sex, batch effect and estimated cell proportions as covariates, with sample ID as a random effect. In total, we identified 11 CpG sites that were significantly (false discovery rate (FDR) < 0.05) changed following BCG vaccination (Figure 2A, Table S2). Among these, five CpG sites showed increased methylation, while six CpG sites displayed decreased methylation 90 days after vaccination. Notably, the changes observed at T14-T0 for eight out of the 11 CpG sites were converse to those observed at T90-T0 (Figure 2B). This observation potentially indicates the presence of distinct epigenetic mechanisms during different time intervals after BCG vaccination. To verify this finding, we further checked the 73 CpG sites with suggestive significance (P<1×10^-5^), and identified consistent patterns (Figure 2C). Pathway analysis of the genes annotated to the 11 CpG sites revealed enrichment in pathways related to Human papillomavirus infections, VEGFA-VEGFR2 signaling and tight junction, which were reported to be related to the immune response to infections previously^25,26^ (Figure S2A).

**Figure 2.**
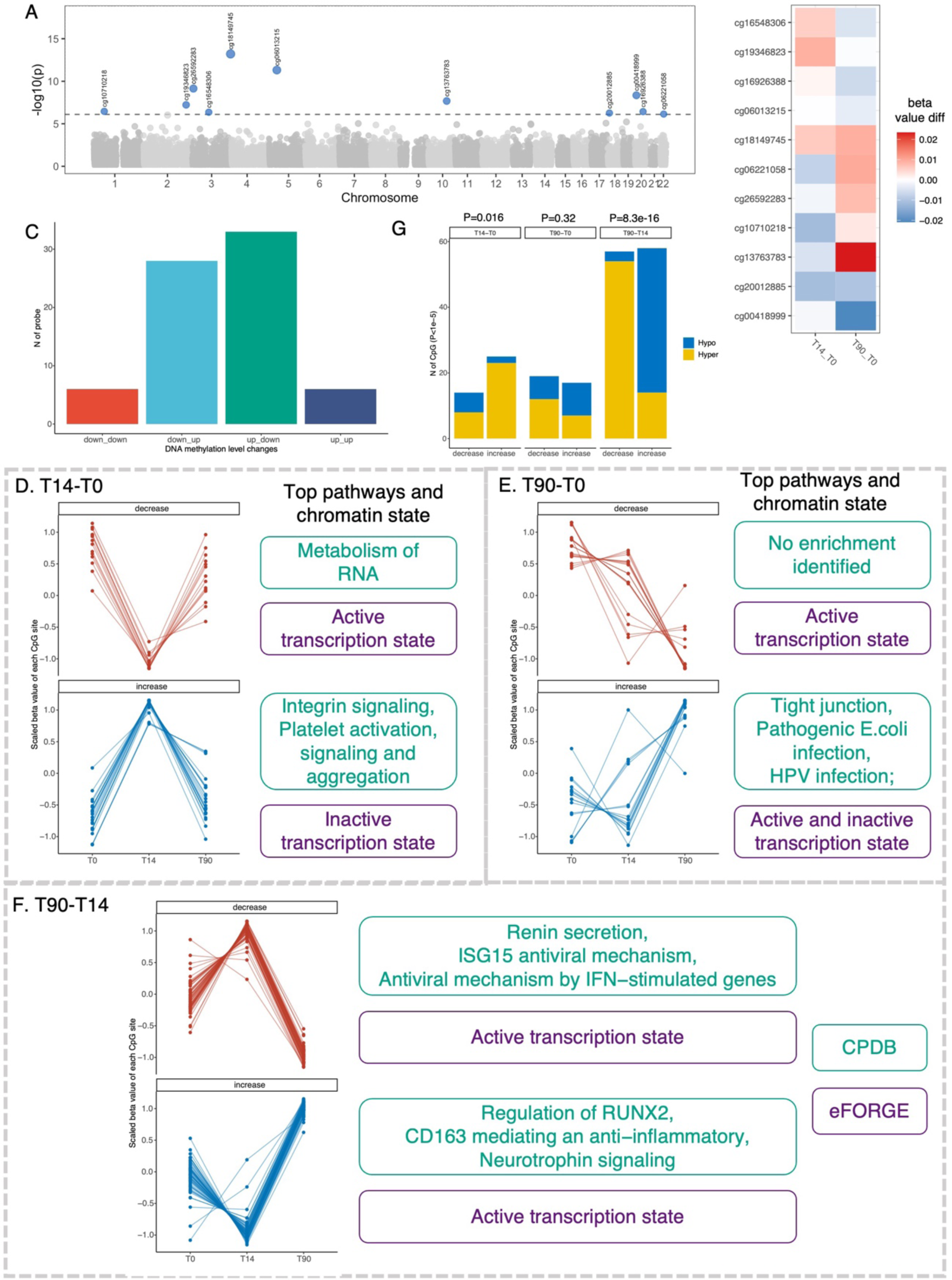
BCG vaccine induced short-term and long-term methylation changes. (A) Manhattan plot showing DNA methylation changes over time after BCG vaccination. Differentially methylated CpG sites (N=11) with false discovery rate (FDR) < 0.05, were highlighted and labeled with the probeID. (B) Heatmap of hierarchical clustering on changes of identified 11 CpG sites at T14 and T90 compared to T0, which are represented as the differences in DNA methylation beta values. (C) Bar plot of the number of CpG sites with significant change over time with P<1×10-5 in different groups. Down_down group indicates DNA methylation was decreased at both T14 and T90 compared with T0, similarly, down_up, decreased at T14 and increased at T90, up_down, increased at T14 and decreased at T90, and up_up, increased at both T14 and T90. (D-F) Change patterns and pathway enrichment analysis of CpG sites identified which assessed the DNA methylation changes at day14 and day0 (T14-T0, D), day90 and day0 (T90-T0, E), as well as day90 and day14 (T90-T14, F) with P<1×10-5. Each panel includes line charts showing the patterns of changes with two plots showing decrease (upper) and increase (lower) change, respectively, and the top enriched pathways and chromatin state were labeled right to each line chart. The pathway enrichment analyses were performed by CPDB (http://cpdb.molgen.mpg.de/) an online tool of gene set analysis, the chromatin enrichment analysis was performed by eFORGE (https://eforge.altiusinstitute.org/). (G) Bar plot showing the number of decreased and increased CpG sites identified from T14-T0, T90-T0 and T90-T14, respectively. Blue represents hypomethylated sites and yellow represents hypermethylated sites. The P value on the top of the figure showing the enrichment hypomethylated/hypermethylated sites in increase/decrease status (Fisher’s exact test).

We next checked the association between the identified CpG sites and other traits using the EWAS catalog (http://www.ewascatalog.org) (Table S3). Five out of the 11 CpG sites have been previously associated with age. Two of these sites were reported to be associated with proteins that involved in B cell development. For instance, cg06221058 (mapped to gene *MYH9*) was reported to be associated with age and IGMH (Immunoglobulin heavy constant Mu) protein levels. cg06013215 (mapped to gene *PTGER4*) was reported to be related to protein amounts of IGLL1 (Immunoglobulin lambda-like polypeptide 1) and FCER2 (Fc Epsilon Receptor II).

Considering the cell-type specificity of DNA methylation markers, the methylation level of these 11 significant CpG sites was then correlated with the estimated cell proportions at each time point (Figure S2B). At all three time points, a strong correlation pattern was observed between the CpG sites and estimated neutrophil proportion, and consistent patterns were identified when correlating the CpG sites with cell counts of neutrophil subtypes measured by flow cytometry (Figure S2C). The top correlated neutrophil subtypes included CD10+ CD66b, CD10+ CD62L, and PDL1-CD62LP neutrophils.

### BCG vaccination induced short-term and long-term changes in DNA methylation

As shown in Figure 2 B-C, the observed patterns of DNA methylation changes suggested the involvement of distinct epigenetic mechanisms in the short- and long-term effects following BCG vaccination. To gain further insights into these effects, we compared the DNA methylation changes between day14 and day0 (T14-T0), day90 and day0 (T90-T0), as well as day90 and day14 (T90-T14) respectively, using mixed effects models. Three CpG sites from the T14-T0 model, five from the T90-T0 model, and sixty-four from the T90-T14 model were identified with genome-wide significant changes (FDR<0.05). In order to capture the overall dynamics of DNA methylation after BCG vaccination, CpG sites reaching suggestive significance threshold (P<1×10^-5^) were also included in the downstream analysis (Table S4-S6).

In the T14-T0 model, which represents the short-term effect of BCG vaccination on methylome, we identified 39 CpG sites (P<1×10^-5^). Among these, 14 CpG sites exhibited decreased methylation at T14 compared to T0, while 25 CpG sites showed increased methylation. Notably, all of these sites demonstrated an opposite direction of change when comparing T90-T14 (Figure 2D, Figure S3A). This suggests that the later time period, from T14 to T90, might represent a recovery phase following BCG vaccination. Using functional enrichment tools experimentally derived Functional element Overlap analysis of ReGions from EWAS(eFORGE)^27^, we found that the demethylated CpG sites were enriched in active transcription states, and the gene mapped to these sites were enriched in pathways involved in the metabolism of RNA, indicating an active transcriptional activity of the demethylated sites at T14. In contrast, the methylated CpG sites were enriched in an inactive transcription state, and were involved in pathways related to integrin signaling and platelet activation and aggregation (Figure 2D, Figure S4D). These results suggested that the short-term DNA methylation alterations induced by BCG vaccination may be involved in the regulation of transcription activity and may return to their original levels over time.

The long-term effect model, T90-T0, revealed 17 methylated CpG sites and 19 demethylated CpG sites (P<1×10^-5^, Figure 2E, Figure S3B) associated with BCG vaccine. These methylated CpG sites exhibited enrichment in both active and inactive transcription states, and the genes mapped to these CpG sites were enriched in pathways such as tight junction and bacterial and viral infection (Figure 2E, Figure S4E). This suggests that the persistent long-term effect of BCG on epigenetics may be related to the response to pathogenic infection. The persistent effect of BCG vaccination on epigenetics at 90 days post-vaccination reflects a lasting epigenetic memory. Interestingly, the majority of the increased CpG sites (64.7%) first slightly decreased at T14 and then increased at T90, indicating that the majority of the increased changes at T90 actually occurred after T14 (Figure 2E, Figure S3B). These results suggested a “late” epigenetic effect of vaccination, in contrast to the “immediate” transcriptional effect of vaccination^13^.

To gain a deeper understanding of the “late” effect of BCG vaccination, we conducted a comparison of methylation profiles between T90 and T14. Through this analysis, we identified 115 CpG sites that displayed suggestively significant changes (P<1×10^-5^). Among these, 58 CpG sites showed increased methylation at T90 compared to T14 after initially being demethylated at T14, surpassing even the levels observed at T0. In contrast, the demethylated CpG sites exhibited the opposite pattern of methylation changes (Figure 2F, Figure S3C). The genes mapped to the identified CpG sites were enriched in pathways related to active transcription state, inflammatory response (e.g., ISG15 antiviral mechanism, antiviral mechanism by IFN-stimulated genes, and CD163 mediated anti-inflammatory) and nervous system (e.g., renin secretion and neurotrophin signaling) (Figure 2F, Figure S4F). Interestingly, we found that the methylated CpG sites identified from comparison of T90 vs T14 were enriched in hypo-methylated sites, while the demethylated CpG sites were enriched in hypermethylated sites (Fisher’s exact test, P=8.31×10^-6^, Figure 2G, Figure S4C), aligning with the concept of epigenetic aging drift^28^. However, the increased CpG sites identified from the T14 vs T0 comparison model were enriched in hyper-methylated sites, contrasting with the pattern observed in comparison of T90 vsT14 (Figure S4 A-B).

### BCG-induced DNA methylation changes were associated with the alteration of plasma inflammatory proteins

We previously reported that BCG vaccination reduced circulating inflammatory markers at both 14 days and 90 days after vaccination^9^. Given the significant changes in DNA methylation profiles upon BCG vaccination observed in this study and that the enriched pathways were related to immune responses to infections, we further investigated whether BCG-induced DNA methylation changes were associated with the altered concentration of circulating inflammatory proteins. In our group’s previous findings, we identified 24 proteins exhibiting significant short-term decreases and 10 proteins that exhibited long-term decreases after BCG vaccination^9^. We correlated these proteins with CpG sites identified in T14 vs T0 (No. CpG =39) and T90 vs T0 (No. CpG = 36) comparison, respectively. We found that the DNA methylation changes in four CpG sites identified from T14 vs T0 comparison were significantly associated with alterations in two proteins upon BCG vaccination, with FDR<0.05 (Table S7). Specifically, two CpG sites were associated with CD6 (cg26624673 near gene *ADHFE1* and *MYBL1*, and cg10813029 near gene *TGIF1* and *DLGAP1*), and two were associated with OPG (cg20782252 near gene *MAD1L1* and *ELFN1*, and cg12598528 near gene *KCNT2*). In addition, we identified 77 CpG-protein pairs that reached nominal significance (P<0.05), among which we observed a consistent pattern in which the CpG sites exhibiting increased methylation at T14 in comparison to T0 were inversely correlated with proteins that exhibited a decrease at T14 relative to T0 (Figure S5A). However, the CpG sites identified from the comparison of T90 vs T0 didn’t show any significant association with protein abundance (FDR < 0.05, Table S7, Figure S5B). These results indicate that BCG-induced DNA methylation changes are associated with reduced concentrations of inflammatory proteins in the circulation, particularly regarding DNA methylation changes from T0 to T14. This suggests that short-term DNA methylation changes may play a role in regulating the reduction of circulating inflammatory proteins induced by BCG vaccination.

### Baseline epigenetic profiles were associated with the trained immunity responses after BCG vaccination

Next, we tested if the epigenetic profiles at baseline were associated with trained immunity (TI) responses, which were defined as the fold changes of cytokine productions stimulated with *S. aureus* at T90 compared to T0 (Figure 1). We measured both the monocyte-derived cytokines (IL-1β, IL-6, and TNF-α), and IFN-γ, which represents the lymphoid cellular response. Our results revealed that 41 CpG sites at baseline were significantly associated with the increased IFN-γ production capacity at T90 compared to T0, denoted as TI (IFN-γ) (FDR<0.05, Figure 3A, Table S8). The strongest association was observed at cg16685860 (P=6.02×10^-10^), located near gene *PLD2*. We identified 22 out of 41 CpG sites that were positively associated with TI (IFN-γ). The genes mapped to these sites were enriched in pathways including phospholipases, Kisspeptin receptor system, and LPA receptor mediated events. Conversely, 19 out of the 41 sites were negatively associated with TI (IFN-γ), and the genes mapped to these sites were enriched in the apoptosis pathway and FAS pathway, which was also related to programmed cell death (Figure S6A). We did not find any significant associations between baseline DNA methylation and TI (IL-1β) or TI (TNF-α), except for one CpG site (cg07586956, mapped to *PCBP1* and *C2orf42* genes) that showed an association with TI (IL-6) (P=5.40×10^-10^). Furthermore, we assessed the association between DNA methylation at 14 and 90 days after BCG vaccination and TI (Table S10-S11). We found a total of 9 CpG sites at T14 and 95 CpG sites at T90 were linked to TI (IFN-γ) (Table S10). For TI (IL-1β), we identified 32 CpG sites at T14 and only three CpG sites at T90 (Table S11), showing different patterns compared to the analysis of TI (IFN-γ), where more sites were identified at T90.

**Figure 3.**
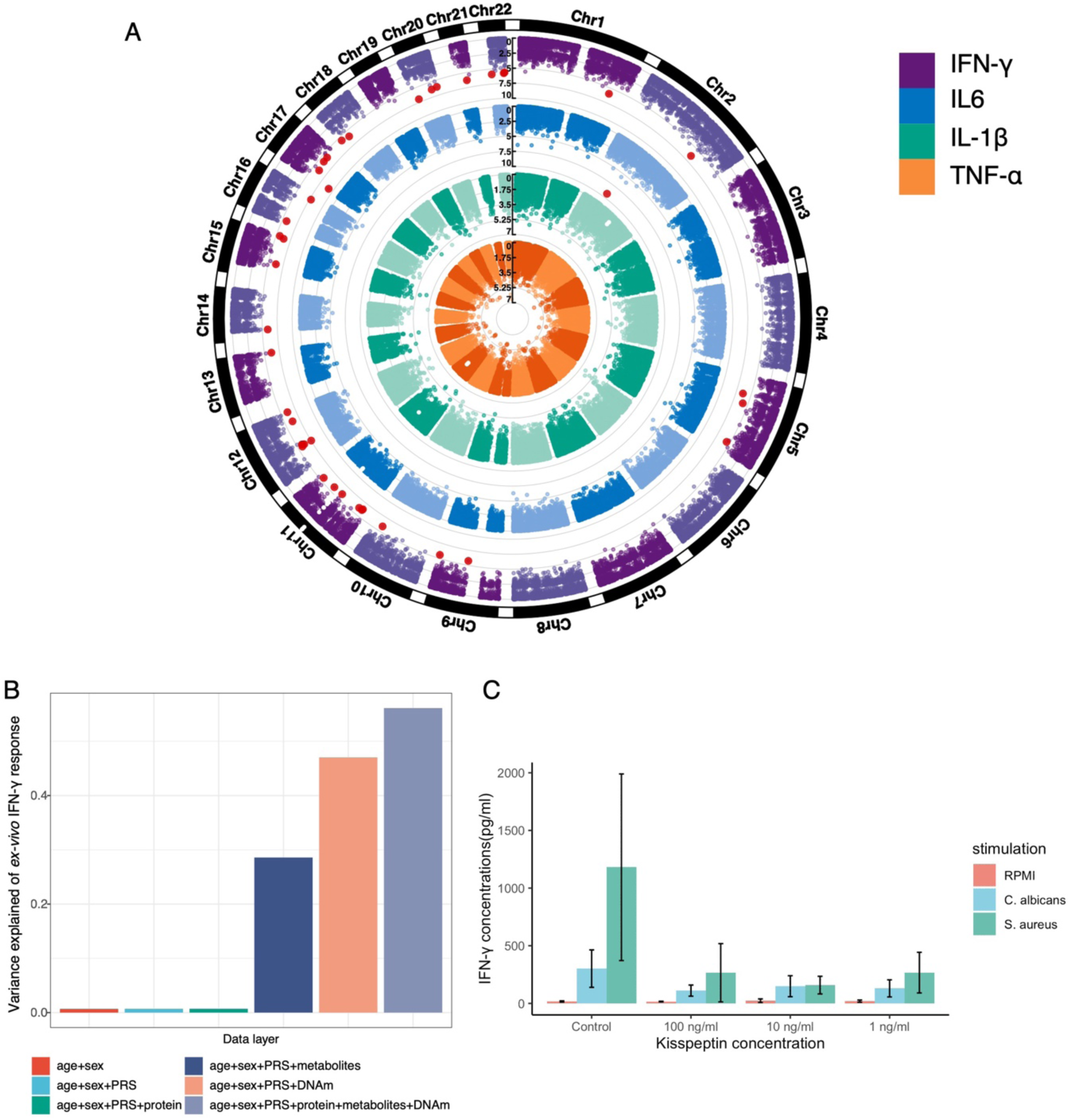
Baseline epigenetic markers and ex-vivo cytokine production changes. (A) Manhattan plot showing the association of baseline DNA methylation level and ex-vivo immune response, represented by fold-change of TNF-α, IL-1β, IL6 and IFN-γ production stimulated by *S-aureus* 90 days after BCG vaccination compared to baseline (from middle to outside). Genome-wide significant CpG sites (FDR<0.05) are highlighted by red. (B) Bar plot showing the variation in *ex vivo* IFN-γ response explained by data from various omics layers, including general information (age and sex), genetics (PRS), baseline DNA methylation, baseline inflammatory proteins, and baseline plasma metabolites. (C) Experimental validation. Chr: chromosome; AUC: area under the curve; CI confidence interval. PRS: polygenic risk score; DNA methylation: DNA methylation; protein: inflammatory proteins.

We further checked the association of the identified CpG sites with other traits by utilizing the EWAS catalog (Table S9). Among the 41 CpG sites, 18 were previously found to be associated with age (Figure S6B). Interestingly, we discovered that five CpG sites (cg03433260 near gene *BID*, cg09391860 near gene *ZNF335* and *MMP9*, cg18591181 near gene *NIM1K*, cg25937862 near gene *MRC2*, and cg22032521 near gene *HYLS1*) showed associations with five protein levels (EBAG9, GZMK, HAPLN4, LYN, and OSM) that have been previously reported in blood.

Then, we investigated how much variance in TI (IFN-γ) could be explained by different omics layers, including polygenetic risk score (PRS)^29^, inflammatory proteins, metabolites, and DNA methylation. Comparing age, sex, and PRS and baseline inflammatory proteins, we discovered that baseline DNA methylation explained the largest proportion of variance of TI (IFN-γ), followed by the baseline metabolites (Figure 3B). A combined model of all data layers explained 56.1% variance for TI (IFN-γ). Further randomization tests confirm the significance of the finding (P value <0.001) (Fig S6D). We also checked if SNPs located proximal to the CpG sites associated with TI (IFN-γ) were enriched in cytokine-QTL of IFN-γ, but we did not identify any significant enrichment (Table S12, Figure S6C).

### Kisspeptin modulates IFNγ production capacity

DNA methylation in the kisspeptin receptor system has been identified to influence IFN-γ responsiveness after BCG vaccination in this study. Kisspeptins are proteins encoded by the *KISS1* gene that have been initially described to inhibit metastases in cancer and induce secretion of gonadotropin-releasing hormone (GnRH), but recently an increasing role in immunomodulatory effects have been described^30^. To functionally validate whether kisspeptins modulate IFN-γ production, human peripheral blood mononuclear cells were incubated for one hour with various concentrations of recombinant kisspeptin-10 (from 1 ng/ml to 100 ng/ml), followed by stimulation with either heat-killed *C. albicans* or *S. aureus* for 7 days. Kisspeptin significantly inhibited IFN-γ production capacity, validating its immunomodulatory role (Figure 3C). In contrast, no effects on IL-17 and IL-22 production, another two cytokines produced by T-cells, were observed (not shown). This demonstrates the inhibitory effect of kisspeptin on IFN-γ production and argues that DNA methylation of *KISS1* likely impacts interferon production.

### BCG-induced DNA methylation changes were associated with *ex vivo* cytokine production changes

In order to investigate if the BCG-induced DNA methylation changes are associated with cytokine production changes, we further performed association analyses between Trained immunity and the differences of DNA methylation levels (DNAm-C) between any two time points (T14-T0, T90-T0, and T90-T14, Table S13). Notably, T90-T0 DNAm-C showed a significant association with trained immunity for all four traits, with the following numbers of significant associations, TNF N=2, IFN-γ N=28, IL-1β N=14 and IL-6 N=3 (FDR<0.05, Figure S7A). This indicates that BCG-induced long-term DNA methylation changes were associated with *ex vivo* cytokine production changes. To better understand the roles of the trained immunity related DNAm-C of T90-T0, we performed the pathway enrichment analyses of the genes mapped to CpG sites with a P value lower than 1×10^-5^ (Figure S8). The trained immunity response as assessed by IL-1β production against a heterologous stimulus from the model T90-T0 was associated with enrichment in changes in DNA methylation for genes involved in Kinesins and Golgi-to-ER retrograde traffic. Kinesins are motor proteins that ensure the transport of cellular cargo, with an emerging role in supporting immune processes as well^31,32^. On the other hand, changes in IFN-γ heterologous production were enriched in changes in DNA methylation of genes involved in the mTOR signaling pathway (well-known to be involved in T-cell activation and trained immunity^33,34^), the VEGFA-VEFGR2 signaling pathway and other immune related pathways (e.g., IL-2 family signaling and IL-18 signaling pathways). These findings provide a further understanding of the functional implications of the DNAm-C in relation to cytokine production changes.

Interestingly, we observed that some of the CpG sites from the association model between Trained immunity and DNAm-C (T90-T0) (TNF-α N=11, IFN-γ N=7, IL-1β N=7, and IL-6 N=5, Figure S7C) exhibited significant changes upon BCG vaccination (P value <0.05). However, the CpG sites from the association analyses between Trained immunity and DNAm-C (T14-T0) or (T90-T14) did not show significant changes upon BCG vaccination. These findings suggest that the BCG-induced long-term DNA methylation changes, rather than short-term changes, may be involved in *ex*-*vivo* cytokine production alteration. Furthermore, it indicates that the development of immune-related epigenetic memory relies on long-term epigenetic changes. We also observed that TI (IFN-γ)-C was associated with both baseline DNA methylation and DNAm-C of T90-T0 with a higher number of identified CpG sites compared to other cytokines. This finding is consistent with previous findings that reported enrichment of genes near CpG sites with long-term BCG effect in IFN related pathways^20^.

### Bidirectional causality between DNA methylation changes and *ex-vivo* cytokine production changes

The underlying causality of the significant association between trained immunity and DNAm-C remains unknown. To investigate this, we utilized the genotype data from 300BCG to infer a potential *in silico* causal relationship between trained immunity and DNA methylation changes (T90-T0) through mediation analysis^35^. We initially identified SNP – trained immunity - DNAm-C groups that exhibited significant association between every two variables and subsequently conducted bidirectional mediation analysis. In the mediation analysis of Direction1, we hypothesized the effects of SNPs on trained immunity were mediated by DNAm-C, and treated DNAm-C as mediator and trained immunity as outcomes. In the analysis of Direction2, trained immunity was treated as the mediator and DNAm-C as outcome (Figure 4A). The majority of significant mediation results were identified for TI(IFN-γ). Specifically, seven CpG sites showed unidirectional causal relationships in Direction1 (P_Direction1_ < 0.05 and P_Direction2_ > 0.05), two CpG sites of unidirectional causal relationships in Direction2 (P_Direction2_ < 0.05 and P_Direction1_ > 0.05), and six CpG sites with bidirectional causal relationships (P_mediation_ < 0.05, Figure 4B, Table S14). In addition, we also identified two CpG sites that mediated the effect of genetic variants on IL-1β changes (Direction1), along with four with bidirectional causal relationships, but none of the CpG sites were significant in Direction2 (Figure 4B, Table S14).

**Figure 4.**
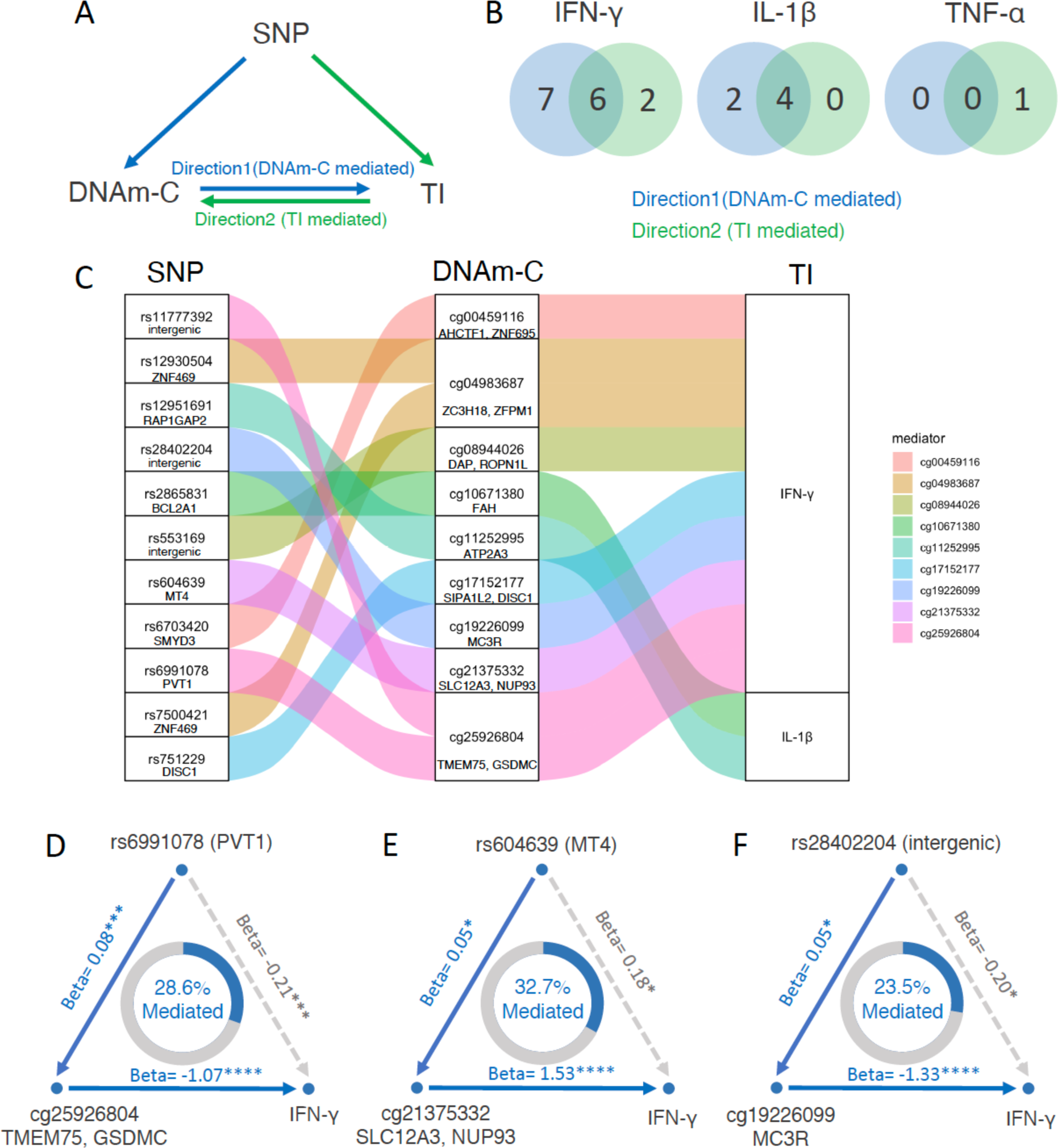
Causal relationship inference by bidirectional mediation analysis. (A) Framework of bidirectional mediation analysis between SNPs, DNA methylation changes (DNAm-C) and cytokine production changes (TI). (B) Number of CpG sites that were significant in mediation results of Direction1 (from DNAm-C to TI), Direction2 (from TI to DNAm-C) and both, for IFN-γ (left) and IL-1β (right). (C) Sankey diagram showing the inferred causal relationship network of Direction1 with mediation P value <0.05. (D-F) Example of causal relationships between SNP, DNAm-C and TI inferred by bidirectional mediation analysis. The beta coefficient and significance are labeled at each edge and the proportions of mediation effect are labeled at the center of ring charts. See also Table S14.

This approach revealed a total of 18 unidirectional mediation linkages: Direction1 analysis (DNAm-C mediated) identified 11 significant linkages involving 9 CpG sites and two cytokines (IFN-γ and IL-1β) (Figure 4C), on the other hand, Direction2 analysis (trained immunity mediated) revealed 7 significant linkages consisted of three CpG sites and two cytokines (IFN-γ and TNF-α). More linkages and CpG sites were identified from Direction1, particularly for IFN-γ and IL-1β, indicating the important role of persistent epigenetic modifications in regulating changes in both monocytes and T-cell derived cytokine production. For instance, cg25926804, located near the *TMEM75* and *GSDMC* gene, mediated 28.6% of the effect of rs6991078 on TI(IFN-γ) (Figure 4D). *GSDMC* was reported to be involved in defense response to bacterium previously^36,37^. Similarly, cg21375332 (near *SLC12A3* and *NUP93*), mediated 32.7% of the effect of rs604639 on IFN-γ changes (Figure 4E). *SLC12A3* is a receptor for the pro-inflammatory cytokine IL-18, and was reported to contribute to IL-18-induced cytokine production, including IFN-γ, IL-6, IL-18 and CCL2^38^. Interestingly, we also found cg19226099, located in *MC3R,* to act as a mediator for IFN-γ (Figure 4F). MC3R is the receptor for melanocyte-stimulating hormone (MSH) and adrenocorticotropic hormone (ACTH) was reported to be related to a delay in the age of puberty onset^39^, suggesting the importance of DNA methylation changes in genes related to hormone that mediating the effect of cytokine production changes after BCG vaccination. In the results from Direction2, where the trained immunity was treated as a mediator, few results were identified compared to Direction1, including cg07420470, located near *RAB3GAP2,* was mediated by TI (IFN-γ) (Figure S9). *RAB3GAP2* is involved in the regulated exocytosis of neurotransmitters and hormones^40^.

### Sex-specificity of BCG effect on DNA methylation and the relationship between DNA methylation and immune responses

In a previous study^9^, sex-specificity in BCG-induced inflammatory protein changes was observed, indicating that the BCG vaccination might act differently in men and women. To further investigate this phenomenon, we stratified our samples by sex and assessed the short- and long-term changes separately in males and females, utilizing the same model as in the previous analysis. Our analysis revealed that following BCG vaccination, there were suggestive significant short-term changes in 18 CpG sites among males, and 31 CpG sites among females (P<1×10^-5^, Table S15) without any overlap, and most of the identified CpG sites show different direction of changes indicating a strong sex-specificity (Figure 5A-B). Specifically, at T14 compared to T0, we observed 13 demethylated sites (72.2%) in males and 10 demethylated sites (32.3%) in females. Through enrichment analysis, we identified sex-specific pathways associated with these methylation changes. Among females, the sex-hormone related pathway GnRH secretion (also modulated by kisspeptins, see above) displayed significant involvement, while among males, functions related to the nervous system, including neurotransmitter receptors and postsynaptic signal transmission, GABA receptor activation, and Renin secretion, were found to be prominent. Additionally, we identified a few shared pathways between both sexes, such as those related to potassium channels (pathway potassium channels in females, and pathway G protein gated potassium channels and Inwardly rectifying K+ channels in males) (Figure 5E).

**Figure 5.**
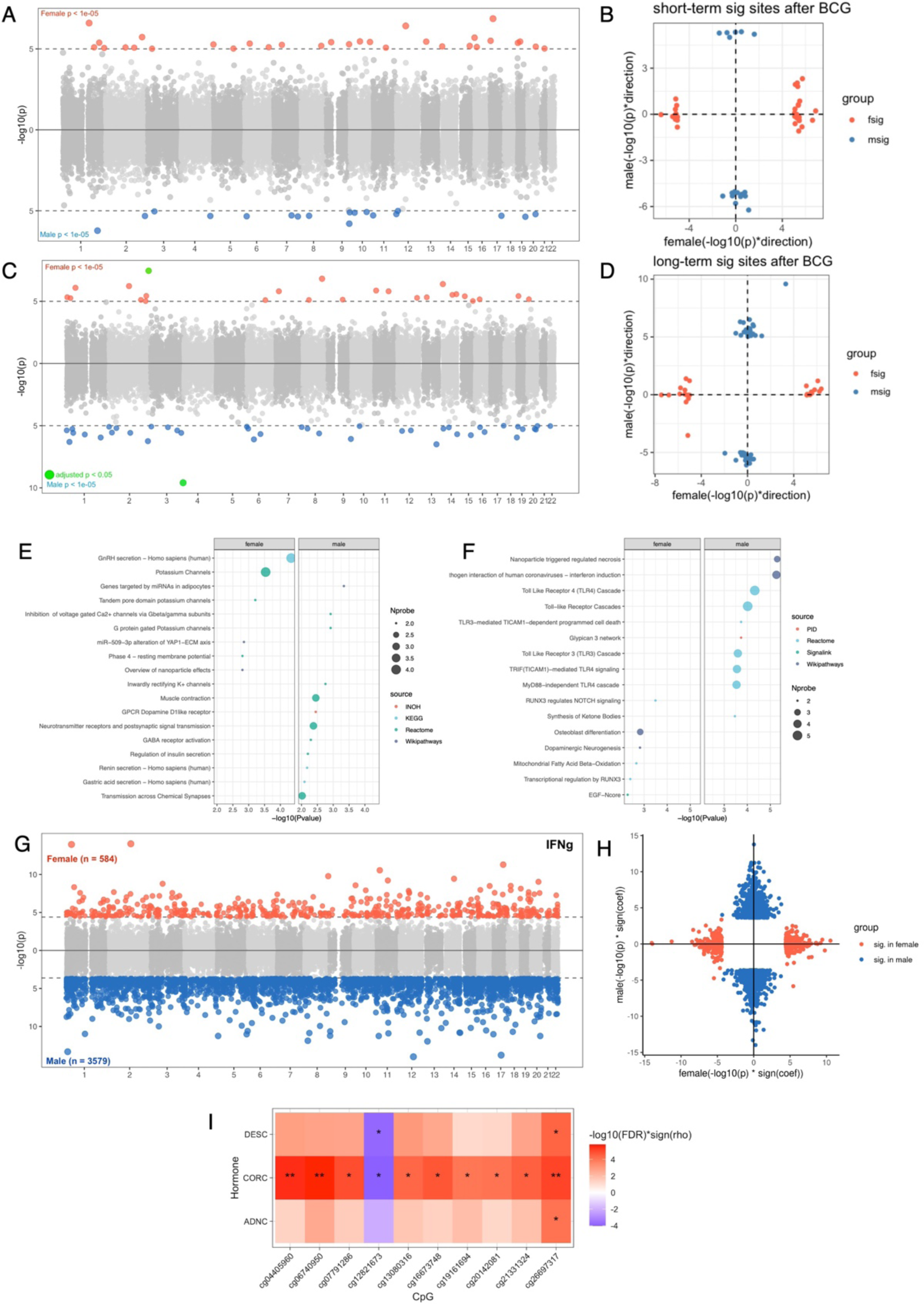
Sex-specific effect on BCG vaccination and the association with ex-vivo cytokine production changes. (A) Miami plot showing the short-term DNA methylation changes upon BCG vaccination (T14-T0) in females (upper) and males (lower). CpG sites with P<1×10^-5^ were highlighted by red (female) and blue (male). (B) Scatter plot showing the consistency of results from males and females regarding the short-term effect. The x-axis represents the -log10 P value with the sign of change direction in females, and the y-axis showing the same value in males. The dots are significant CpG sites identified from female (red dots) and male (blue dots) respectively with P<1×10^-5^. (C) Miami plot showing the long-term DNA methylation changes upon BCG vaccination (T90-T0) in females (upper) and males (lower), the only CpG site passed FDR significance was highlighted by green. (D) Scatter plot showing the consistency of results from males and females regarding the long-term effect. (E-F) Dot plot showing the pathway enrichment of genes annotated to the identified CpG sites in short-term (E) and long-term (F), with two panels for females (left) and males (left) respectively. (G) Miami plot showing the association between baseline DNA methylation and *ex vivo* production changes of IFN-γ in females (upper) and males (lower) with FDR<0.05. (H) Scatter plot showing the consistency of the results from G in males and females. The x-axis represents the -log10 P value with the sign of correlation coefficient in females, and the y-axis showing the same value in males. (I) Heatmap showing the spearman correlation between TI (IFN-γ) associated CpG sites and baseline Hormone levels of androstenedione (ADNC), cortisol (CORC) and 11 deoxy cortisol (DESC). Only significant correlations (FDR<0.05) are shown in this heatmap. Cell colors indicate the -log10(P) with the sign of correlation coefficient of spearman correlation (rho), and the asterisks indicate the significance of the correlation (* FDR<0.05, ** FDR<0.01).

We also identified suggestive significant changes in 42 CpG sites in males, and 25 CpG sites in females as long-term effects after vaccination (P<1×10^-5^, Figure 5C-D, Table S16). Unlike the short-term BCG effect sites, the long-term changes in CpG sites exhibited the most consistent increases or decreases in males, whereas no consistent patterns were identified in females (Figure S10). Pathway analysis revealed that the CpG sites showing long-term effects in males were enriched in functions related to infection and immune responses (host−pathogen interaction of human coronaviruses and Toll−like Receptor pathways) and cell death (nanoparticle triggered regulated necrosis). In contrast, the long-term sites in females were enriched in pathways including RUNX3 regulates NOTCH signaling, dopaminergic neurogenesis, and osteoblast differentiation (Figure 5F). These findings indicate that after BCG vaccination, the DNA methylation profiles undergo different trajectories in males and females, and these BCG-induced methylation changes are associated with distinct functional pathways. In summary, the short-term changes in methylation following BCG vaccination were enriched to functions related to sex hormones in females and the nervous system in males, while the long-term changes were associated with pathways related to pathogenic immune responses in males and the nervous systems in females.

The *ex vivo* cytokine production changes were reported to be associated with inflammatory proteins in a sex-dependent manner^9^. We also observed the sex-specific association between trained immunity of four cytokines and baseline epigenetic profiles by sex-stratified analysis, with a stronger association identified in males than females for all four cytokines tested in this study. In males, we identified 3579 CpG sites at baseline that were significantly associated with TI (IFN-γ) males, whereas only 584 sites showed significant association in females, without any overlap (FDR<0.05, Figure 5G, Table S17). Furthermore, the direction of 1883 out of the total 4161 (45.3%) associations was inconsistent between males and females (Figure 5H). Similar trends were observed for other cytokine markers, such as IL-1β (1107 in males and 62 in females), IL-6 (853 in males and 67 in females), and TNF-α (513 in males and 9 in females) (Table S18). Importantly, these associated sites were not enriched to the CpG sites that were significantly associated with sex at baseline (Fisher’s exact test P > 0.05, Table S19). Instead, pathway analysis revealed that CpG sites associated with TI (IFN-γ) in both males and females were enriched in signal transduction pathways. However, other enriched pathways were specific to males or females. For instance, the top pathways in males included signaling by receptor tyrosine kinases, the Rap1 signaling pathway, and pathways in cancer, while in females, the top pathways were the Hippo signaling pathway, dopaminergic neurogenesis, and NrCAM interactions (Figure S11). To gain insight into the sex specificity of these associations, we also associated the identified CpG sites with five types of sex hormones including androstenedione, cortisol, 11 deoxy cortisol, 17 hydroxy progesterone, and testosterone. Interestingly, 10 CpG sites associated with TI (IFN-γ) in females were strongly associated with cortisol, and two CpG sites were also associated with 11 deoxy cortisol (spearman correlation FDR <0.05, Figure 5I), while for the sites identified in males, we did not identify any significant links with sex-hormones.

### Replication of EWAS results

We conducted a replication study to validate our findings using an independent cohort. In this study, 17 individuals were vaccinated with BCG, and blood samples were collected before vaccination (T0) and 3 months (T90) after vaccination followed by DNA methylation profiling. The assessment of *ex vivo* cytokine changes followed was the same protocol as the discovery cohort. Firstly, we compared the difference of DNA methylation levels of 36 CpG (P<1×10^-5^ in the discovery cohort) between T90 and T0 in the replication cohort, and we identified that 21 out of 36 CpG sites showed the same direction, and two out of the 21 CpG sites showed nominal significance (P<0.05) (Table S20). Due to the limited number of male samples collected available (N=4), we were only able to test the female-specific results. We tested the long-term female specific CpG sites (N=25), and 12 out of 25 CpG sites showed consistent directions of changes with one CpG site reaching nominal significance (P<0.05, Table S21).

Next, we replicated the associations between baseline DNA methylation and TI (IFN-γ), using the same model used in the discovery cohort. Out of 40 CpG sites, we identified five CpG sites that could be replicated with nominal significance (P<0.05) and consistent direction of effect (Table S22). Additionally, for the female-specific associations, we found that 34 out of 582 CpG sites can be replicated with nominal significance (P<0.05) and consistent directions of effect. Among these, 10 CpG sites reached the FDR significance (FDR<0.05, Table S23).

## Discussion

Epigenetic reprogramming has been proposed as a mechanism underlying BCG-induced innate immune memory. In this study, we provide evidence for the dynamic landscape of DNA methylation changes following BCG vaccination and elucidate the role of DNA methylation in immunological responses induced by BCG vaccination. Our findings demonstrate that BCG induces persistent immune-related DNA methylation changes for up to at least 3 months, likely reflecting epigenetic memory at the DNA methylation level (Figure 6).

**Figure 6.**
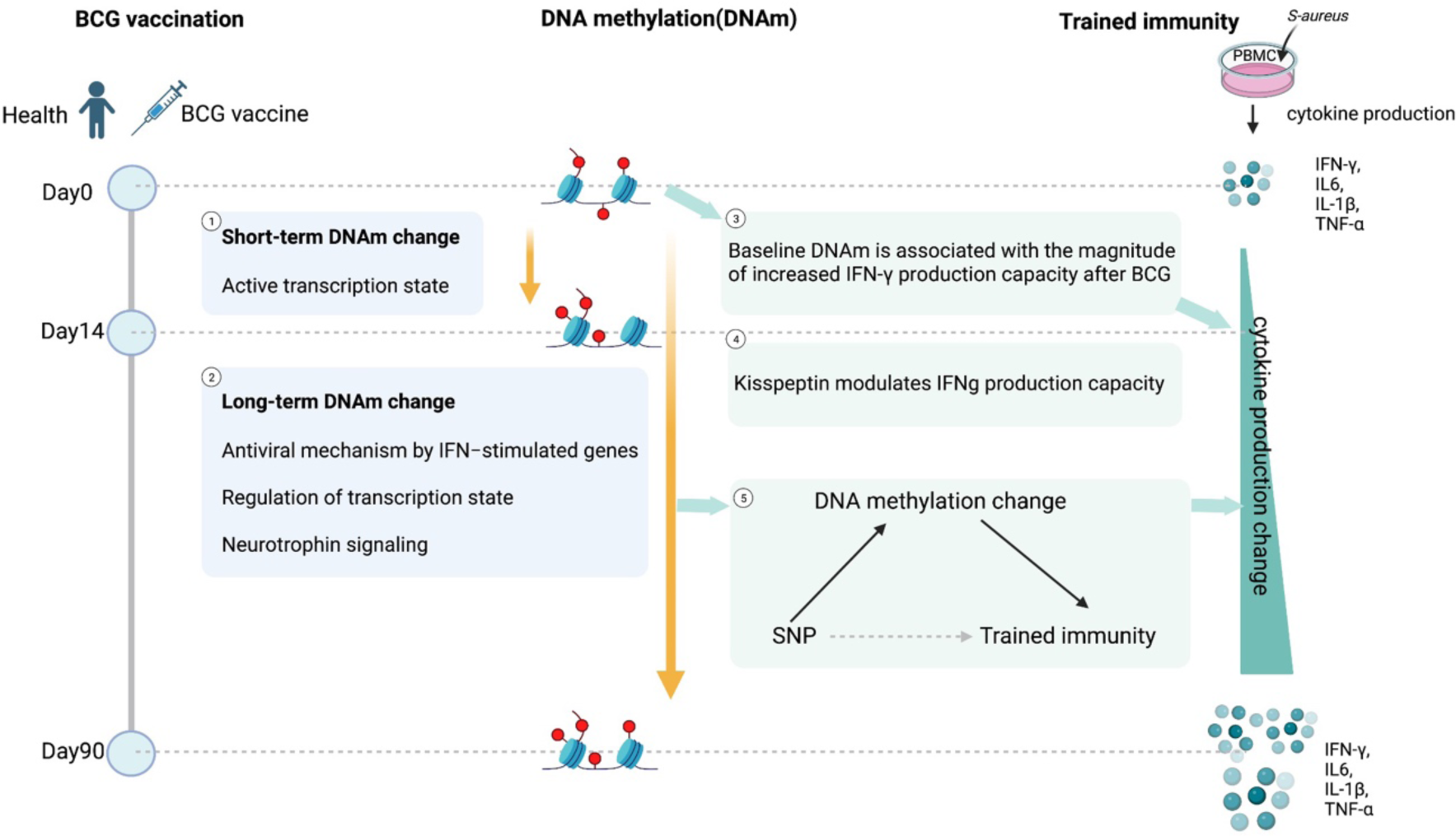
Summary of findings of this study.

While long-term epigenetic processes have been proposed earlier to mediate trained immunity, most studies to date have investigated histone modifications^41,42^. A recent study investigated changes in DNA methylation in monocytes of young children^20^, while we suggested that DNA methylation changes after BCG vaccination in adults as well^22^. A comprehensive analysis of the impact of DNA methylation on innate immune memory in adults was missing. In this study, we observed distinct patterns of short- and long-term DNA methylation changes following BCG vaccination. Previous research has outlined distinct stages of epigenetic reprogramming in the induction of trained immunity, involving acute stimulation of innate immune cells leading to active transcription of proinflammatory factors, followed by a resting stage, where epigenomic changes are partially reversed after stimulus cessation^43^. In our study, we observed similar acute effects on DNA methylation at day 14 that returned back to baseline by day 90 following BCG vaccination. Notably, the demethylated CpG sites associated with this acute effect were enriched in an active transcription state, consistent with the previous finding that BCG vaccination induced acute and resting stages. Besides, persistent epigenetic changes, potentially contributing to epigenetic memory, were enriched in infection-related pathways and associated with *ex vivo* cytokine changes that represented innate immune memory. These findings suggest that epigenetic memory partly underlies the induction of immune memory. Furthermore, we identified a “late” epigenetic reprograming that includes decreased hypermethylation and increased hypomethylation. DNA methylation is a known stable biomarker of aging, with a tendency towards increased hypermethylation in aging processes^44^. Interestingly, our results suggest that the “late” effect of BCG vaccination opposes the DNA methylation changes associated with natural aging, indicating a distinct epigenetic influence of BCG vaccination.

BCG vaccination has been shown to induce epigenetic reprogramming, such as DNA methylation^45^ and histone modifications^2^, which play a critical role in trained immunity or immunological memory. In our study, we investigated the association between DNA methylation levels and immunological memory following BCG vaccination, specifically focusing on *ex vivo* cytokine production changes of four cytokines upon *S. aureus* stimulation. We found that baseline DNA methylation explained a significant proportion of variance of T-cell heterologous IFN-γ responses and this may also play a role in the process of immune memory^46^. These results indicated that individual differences in long-term innate immune effects induced by vaccines depend on intrinsic DNA methylation levels prior to vaccination, and DNA methylation status at certain CpG sites may facilitate the immune responses after BCG vaccination. Among the pathways modulated by DNA methylation that was found to strongly modulate heterologous immune responses, we identified baseline CpG sites enriched in the kisspeptin pathway. Kisspeptins are peptides that induce the release of GnRH, but with an increasing number of studies showing immunomodulatory effects^47,48^. Indeed, functional validation experiments demonstrate their modulatory effects on IFNγ production, strengthening the hypothesis of their impact on long-term immune responses.

Moreover, we identified that BCG-induced DNA methylation changes at day 90 compared to baseline, and these were associated with *ex vivo* cytokine changes including both IFN-γ and IL-1β, representing responses from both monocytes and T-cell derived cytokine responses. Previous studies have shown that the stimulation of innate immune cells can leave an “epigenetic scar”, making the cells more responsive to a different stimulation^43^. Our results suggest that epigenetic memory is represented by a combination of innate immune training and adaptive immune memory^13,49^. Furthermore, through mediation analysis, we showed the potential *in-silico* causal relationship between BCG induced persistent methylation changes and trained immunity. We identified more CpG sites that may mediate the regulation of genetic variants on IFN-γ and IL-1β changes after BCG vaccination, suggesting the DNA methylation acts as a modulator of immune response. The identified CpG sites were located in genes related to pathogenic responses and cytokine productions, which stressed the importance of epigenetic reprogramming at CpG sites on the regulation of trained immunity. For example, the DNA methylation changes of cg21375332 near *SLC12A3* gene mediated the regulation of genetic variants on IFN-γ changes, and this was consistent with previous study that *SLC12A3* was involved in IL-18-induced IFN-γ production^38^. We also revealed that CpG site near *GSDMC*, a gene involved in the defense response to bacterium, mediated the changes of IFN-γ after BCG vaccination.

Several studies have demonstrated that the BCG vaccination can exhibit sex-specific effects^50,51^. In a study investigating the protective effects of neonatal BCG vaccination, it was observed that boys exhibited strong protective effects within the first week after vaccination, which diminished thereafter. Conversely, girls showed weaker protection initially but demonstrated stronger effects later on^51^. In our study involving adult participants, we also identified sex-specific DNA methylation changes induced by BCG vaccination, both in the short and long term. Short-term DNA methylation changes in females were enriched to the GnRH pathway, which influences hormone secretion and immune system development^52^. Long-term effects in males involved DNA methylation changes related to Toll-like receptors (TLRs), particularly TLR4, which recognizes bacteria lipopolysaccharide (LPS) and activates innate immunity. Conversely, long-term effects in females were observed in CpG sites associated with neuronal functions, including Notch signaling and dopaminergic neurogenesis. It is worth noting that the neonatal BCG vaccination was reported to improve neurogenesis in mice^53^.

Moreover, baseline DNA methylation levels impact the cytokine changes following BCG vaccination in a sex-specific manner. We identified the dopaminergic neurogenesis pathway in females to influence the IFN-γ associated CpG sites, which confirmed the importance of epigenetic modifications in the neuronal crosstalk with the immune system after BCG vaccination in females. We also revealed that the IFN-γ associated CpG sites in males were enriched in the receptor tyrosine kinases pathway, which was known as key regulators of an uncontrolled immune response and played a critical role in the control of autoimmune disorders^54^. Interestingly, we observed an association between IFN-γ associated CpG sites and cortisol, specifically in females. Cortisol, a mediator of stress-induced immune-suppression, may affect both innate and adaptive immunity. A previous study revealed that infants with higher cortisol response to pain and lower delayed-type hypersensitivity response to BCG vaccination^55^.

This study has also some limitations that should be acknowledged. Firstly, it is important to note that DNA methylation is cell-type specific and cannot be assessed at single-cell resolution in this study. Secondly, the DNA methylation analysis conducted using the EPIC array only covers approximately 3% of the CpG sites in the genome. Finally, although we were able to partially replicate our results in an independent cohort, the validation study had limited statistical power due to the limited sample size.

In conclusion, our study provides valuable insights into the dynamic changes of DNA methylation following BCG vaccination and highlights the induction of immune-related epigenetic memory through these dynamic DNA methylation changes. The observed DNA methylation changes play a regulatory role in immunological responses triggered by BCG vaccination, underscoring the presence of immune-related epigenetic memory.

## Method

### Study design and cohort description

In the 300BCG study, 303 healthy Dutch individuals were included from April 2017 until June 2018. After obtaining the written informed consent, blood was collected, followed by the administration of a standard dose of 0.1 mL BCG (BCG-Bulgaria, InterVax) intradermally in the left upper arm by a medical doctor. Two weeks and 3 months after BCG vaccination, additional blood samples were collected and a questionnaire was completed. Exclusion criteria were the use of systemic medication other than oral contraceptives or acetaminophen, use of antibiotics 3 months before inclusion, previous BCG vaccination, history of tuberculosis, any febrile illness 4 weeks before participation, any vaccination 3 months before participation, or a medical history of immunodeficiency. This study was approved by the Arnhem-Nijmegen Medical Ethical Committee (NL58553.091.16).

### DNA methylation measurement and quality control

DNA was isolated from whole blood using a QIAamp blood kit (Qiagen Benelux BV, Venlo, the Netherlands) kit, and the concentration was determined using a NanoDrop spectrophotometer at 260 nm. Genome-wide DNA methylation profiles were measured by Infiniumγ MethylationEPIC array (∼850,000 CpG sites). DNA methylation data were pre-processed in R (version 4.0) with the Bioconductor package Minfi^56^, using the original IDAT files extracted from the HiScanSQ scanner. We removed the samples which had bad call rate(n=4), sex-mismatched (n=1), and we checked whether had mixed-up samples by inspecting the correlation of the beta value for SNPs. Mismatched samples were replaced by a new label with a correlation of larger than 0.7. Quality control was performed to filter bad quality probes with a detection P-value>0.01, cross-reactive probes, polymorphic probes^57,58^, and probes in the sex chromosome. We subsequently implemented stratified quantile normalization^59^. Based on methylation value, cell proportion was estimated using Housman’s method^24^. After quality control and matching with phenotype data, 284 samples and 751,564 probes remained for further analyses. Methylation levels (beta values, β) were converted into M values (log2(β/1-β)), which were used in downstream differential methylation analyses. Extreme outliers in the methylation data were identified using the Tukey method (<1st quartile−3 × IQR; >3rd quartile+3 × IQR) and set as missing.

### Assessment of ex vivo cytokine responses

Peripheral blood mononuclear cells (PBMCs) were isolated from EDTA whole blood with Ficoll-Paque (GE Healthcare) density gradient separation. Cells were washed twice in PBS and suspended in RPMI culture medium (Roswell Park Memorial Institute medium, Invitrogen, CA, USA) supplemented with 50 mg/mL gentamicin (Centrafarm), 2 mM glutamax (GIBCO), and 1 mM pyruvate (GIBCO). Subsequently, the PBMCs were stimulated *ex vivo* with 10^6^ CFU/mL heat-killed *Staphylococcus aureus* or left unstimulated. Cytokine production of TNF-α, IL-6, and IL-1β was measured in 24-hour supernatants, and IFN-γ was measured in supernatants 7 days after stimulation using ELISA. Then, we calculated the fold change in cytokine production (3 months after vaccination compared to baseline). After log10 transformation and correcting for batch effect, the value was used as a measurement of the magnitude of the trained immunity response. For the *in-vitro* validation of the immunomodulatory role of kisspeptin, a similar 7-days stimulation assay for the production of IFN-γ was employed in human PBMCs.

### Association of DNA methylation with BCG effect and immune responses

Principle component analysis (PCA) was performed using DNA methylation data from each time point respectively. The top 30 PCs were associated with variables including age, sex, batch, BMI, smoking, and estimated cell counts by linear regression model.

To assess the DNA methylation changes induced by BCG vaccination, a linear mix effect model was performed for each CpG site, with time point as the independent variable, subject ID as the random effect, and age, sex, batch, and estimated cell counts as the covariates. Besides the overall changes, we also assessed the changes from T0 to T14, T0 to T90, and T14 to T90, using the same model which only included two time points. EWAS of trained *ex vivo* cytokine response at each time point was performed by fitting the robust linear regression model adjusting for age, sex, batch, and estimated cell counts. The *ex vivo* cytokine response was normalized by the inverse rank method. Besides, we also assessed the association between DNA methylation changes and *ex vivo* cytokine production changes using the robust linear regression model. Considering the different estimated cell counts at different time points, we first regress out the cell counts and batch at each time point from DNA methylation M values, and the residuals were obtained. Then we used a robust linear regression model to correlate the DNA methylation changes (differences in the residuals at any two time points T90-T0, T14-T0, and T90-T14), to the trained immunity response with age and gender as covariables. CpG sites passed the threshold of FDR<0.05 were considered significant findings. The sex-stratified analysis was performed in male and female respectively using the same models mentioned above.

The identified CpG sites were annotated to nearby genes by the GREAT annotation tool^60^. A tool called experimentally derived Functional element Overlap analysis of ReGions from EWAS (eFORGE) 2.0^27^ was used to annotate the CpG sites associated with the BCG effect to further understand the function of these CpG sites in transcription activity. We applied this tool to methylated and demethylated CpG sites with P value < 10^-5^ respectively of the overall BCG model. Pathway analysis of genes mapped to identified CpG lists was performed with the online tool CPDB http://cpdb.molgen.mpg.de/. DNA methylation levels and estimated cell proportions at different time points (for 11 CpG sites associated with the overall BCG effect) were performed by Spearman correlation. Neutrophil cell counts were also measured by flow cytometry where the sub-cell types were also measured. And then the cell counts were correlated to 11 CpG sites by Spearman correlation.

The assessment and quality control steps for inflammatory protein data were published previously^9^. Proteins that were significantly decreased at T14 compared to T0 (N=24), and T90 compared to T0 (N=10) were selected based on the previous study^9^. CpG sites that were significantly changed at T14 compared to T0 (N=39), and T90 compared to T0 (N=36) were selected by previous steps in this study. The associations between DNA methylation changes (T14-T0 and T90-T0) and inflammatory protein changes (fold change T14/T0 and T90/T0) were performed by Spearman correlation.

### Variance explanation

We estimated the variance explained by age, sex, PRS, as well as DNA methylation, inflammatory proteins, and circulating metabolites at baseline, on *ex vivo* IFN-γ response. The PRS was calculated from SNPs associated with the trained immune response. Inflammatory proteins at baseline were assessed by Olink inflammation panel^9^. Baseline metabolites were measured and annotated by the General Metabolics (Zurich, Switzerland) using flow injection time-of-flight mass (flow-injection TOF-M) spectrometry^10^.

We first select the features by associating the features in each data level to TI (IFN-γ). If a feature showed a significant association (Spearman correlation, P value <0.05), the feature was included as the candidate predictor. For DNA methylation we included the 41 CpG sites identified by EWAS analysis. For metabolites data, we included 34 metabolites that were significantly associated with TI (IFN-γ, P<0.05). And for protein data, we do not identify any significant proteins that were associated with TI (IFN-γ). Each candidate predictor from each data layer was correlated to other predictors within the data layer to identify collinearity among these predictors. If the features within this layer showed an association (Spearman correlation > 0.4), the feature which showed the least association (based on the p-value) to TI (IFN-γ) was further removed from the candidate predictors. This yielded a unique set of predictors from each layer, which was then used to fit a multivariate linear model to estimate the variance explained by these features for TI (IFN-γ). Finally, 31 CpG sites and 23 metabolites were selected by these approaches and were used in the variance calculation. To account for the inflation that adding more predictors has on the explained variation, the adjusted R^2^ was used as the measure of explained variance. Additionally, we also randomly selected the same number of features (n=73, the number of proteins available in this study) 1000 times, and repeated the approaches mentioned above to get the variance explained by randomly selected features.

### Cytokine-QTL and enrichment analysis

The association between SNP dosages and IFN-γ responses (cytokine QTL) was done using Matrix eQTL R package^61^ with the inversed rank normalized IFN-γ responses value as the dependent factor, adjusting for age and sex. Using the identified IFN-γ associated CpG sites list (N=41, Table S13), cytokine QTL p-values of those SNPs located 250 kb upstream or downstream of those CpG sites were extracted and plotted against random cytokine QTL p-values. By this, the *cis* effect of CpG sites on cytokine QTL was tested.

### Bi-directional mediation analysis

Matrix eQTL^61^ was used to identify the significant pairs of TI-SNPs and DNAm-C SNPs after regressing out age and gender. For SNPs significantly associated with both TI and DNAm-C (P < 0.05), we carried out bi-directional mediation analysis (y = x + m + ε, where y is the outcome, x is the SNP dosage and m represents the mediator) using mediation R package (ref mediation: R Package for Causal Mediation Analysis) to infer the mediation effect of TI or DNAm-C for genetic impacts.

### Replication cohort

We replicated our results in a cohort consisting of 17 individuals from BCG booster cohort^23^. This randomized placebo-controlled trial originally compares different BCG vaccination regimens for identifying their efficacy to establish trained immunity. The 17 volunteers that received one dose of BCG as positive control were used as validation cohorts in the current study. Vaccination was performed intradermally in the left upper arm as the same manner as with the discovery cohort. Blood was drawn at baseline, and three months after the vaccination. The trial protocol was registered under NL58219.091.16 in the Dutch trial registry, and was approved in 2019 by the Arnhem-Nijmegen Ethics Committee. All experiments were conducted in accordance with the Declaration of Helsinki and no adverse events were recorded.

### Data and code availability

DNA methylation data have been deposited at the European Genome-phenome Archive (EGA), which is hosted by the EBI and the CRG, under accession number EGAS00001007498. Code generated to process the data are freely available on Github (https://github.com/CiiM-Bioinformatics-group/BCG_methylation_project)

## Data Availability

All data in the present work have been deposited at the European Genome-phenome Archive (EGA), which is hosted by the EBI and the CRG, under accession number EGAS00001007498.

## Author Contributions

MGN, CX and YL conceptualized and designed the study. CQ and ZL performed the data analysis supervised by YL, CX and MGN. SJCFMM and VACMK recruited the participants and collected the biological material. GK, ASS and PAD performed DNA isolation and supported the functional experiments; helped with participant recruitment and interpretation of the data, with support from AP, LCJdeB, VPM and LABJ. YAM, WL, MG, and AA helped with part of the data analysis. CQ, ZL, CX and MGN wrote the manuscript with input from all the authors. All authors reviewed and approved the manuscript.

## Acknowledgments

YL was supported by an ERC starting Grant (948207) and a Radboud University Medical Centre Hypatia Grant (2018). CJX was supported by Helmholtz Initiative and Networking Fund (1800167) and Deutsche Forschungsgemeinschaft (DFG) Fund (497673685). MGN was supported by an ERC Advanced Grant (833247) and a Spinoza Grant of the Netherlands Organization for Scientific Research.

## References

1. Netea, M.G., Quintin, J., and van der Meer, J.W.M. (2011). Trained immunity: a memory for innate host defense. Cell Host Microbe 9, 355–361. 10.1016/j.chom.2011.04.006.

2. Kleinnijenhuis, J., Quintin, J., Preijers, F., Joosten, L.A.B., Ifrim, D.C., Saeed, S., Jacobs, C., van Loenhout, J., de Jong, D., Stunnenberg, H.G., et al. (2012). Bacille Calmette-Guerin induces NOD2-dependent nonspecific protection from reinfection via epigenetic reprogramming of monocytes. Proc Natl Acad Sci U S A 109, 17537–17542. 10.1073/pnas.1202870109.

3. Arts, R.J.W., Moorlag, S.J.C.F.M., Novakovic, B., Li, Y., Wang, S.-Y., Oosting, M., Kumar, V., Xavier, R.J., Wijmenga, C., Joosten, L.A.B., et al. (2018). BCG Vaccination Protects against Experimental Viral Infection in Humans through the Induction of Cytokines Associated with Trained Immunity. Cell Host Microbe 23, 89–100.e5. 10.1016/j.chom.2017.12.010.

4. Giamarellos-Bourboulis, E.J., Tsilika, M., Moorlag, S., Antonakos, N., Kotsaki, A., Domínguez-Andrés, J., Kyriazopoulou, E., Gkavogianni, T., Adami, M.-E., Damoraki, G., et al. (2020). Activate: Randomized Clinical Trial of BCG Vaccination against Infection in the Elderly. Cell 183, 315–323.e9. 10.1016/j.cell.2020.08.051.

5. Kaufmann, E., Khan, N., Tran, K.A., Ulndreaj, A., Pernet, E., Fontes, G., Lupien, A., Desmeules, P., McIntosh, F., Abow, A., et al. (2022). BCG vaccination provides protection against IAV but not SARS-CoV-2. Cell Rep 38, 110502. 10.1016/j.celrep.2022.110502.

6. Moorlag, S.J.C.F.M., Rodriguez-Rosales, Y.A., Gillard, J., Fanucchi, S., Theunissen, K., Novakovic, B., de Bont, C.M., Negishi, Y., Fok, E.T., Kalafati, L., et al. (2020). BCG Vaccination Induces Long-Term Functional Reprogramming of Human Neutrophils. Cell Rep 33, 108387. 10.1016/j.celrep.2020.108387.

7. Netea, M.G., Joosten, L.A.B., Latz, E., Mills, K.H.G., Natoli, G., Stunnenberg, H.G., O’Neill, L.A.J., and Xavier, R.J. (2016). Trained immunity: A program of innate immune memory in health and disease. Science 352, aaf1098. 10.1126/science.aaf1098.

8. Divangahi, M., Aaby, P., Khader, S.A., Barreiro, L.B., Bekkering, S., Chavakis, T., van Crevel, R., Curtis, N., DiNardo, A.R., Dominguez-Andres, J., et al. (2021). Trained immunity, tolerance, priming and differentiation: distinct immunological processes. Nat Immunol 22, 2–6. 10.1038/s41590-020-00845-6.

9. Koeken, V.A., de Bree, L.C.J., Mourits, V.P., Moorlag, S.J., Walk, J., Cirovic, B., Arts, R.J., Jaeger, M., Dijkstra, H., Lemmers, H., et al. (2020). BCG vaccination in humans inhibits systemic inflammation in a sex-dependent manner. J Clin Invest 130, 5591–5602. 10.1172/JCI133935.

10. Koeken, V.A.C.M., Qi, C., Mourits, V.P., de Bree, L.C.J., Moorlag, S.J.C.F.M., Sonawane, V., Lemmers, H., Dijkstra, H., Joosten, L.A.B., van Laarhoven, A., et al. (2022). Plasma metabolome predicts trained immunity responses after antituberculosis BCG vaccination. PLoS Biol 20, e3001765. 10.1371/journal.pbio.3001765.

11. Kong, L., Moorlag, S.J.C.F.M., Lefkovith, A., Li, B., Matzaraki, V., van Emst, L., Kang, H.A., Latorre, I., Jaeger, M., Joosten, L.A.B., et al. (2021). Single-cell transcriptomic profiles reveal changes associated with BCG-induced trained immunity and protective effects in circulating monocytes. Cell Rep 37, 110028. 10.1016/j.celrep.2021.110028.

12. Zhang, B., Moorlag, S.J., Dominguez-Andres, J., Bulut, Ö., Kilic, G., Liu, Z., van Crevel, R., Xu, C.-J., Joosten, L.A., Netea, M.G., et al. (2022). Single-cell RNA sequencing reveals induction of distinct trained-immunity programs in human monocytes. J Clin Invest 132, e147719. 10.1172/JCI147719.

13. Li, W., Moorlag, S.J.C.F.M., Koeken, V.A.C.M., Röring, R.J., de Bree, L.C.J., Mourits, V.P., Gupta, M.K., Zhang, B., Fu, J., Zhang, Z., et al. (2023). A single-cell view on host immune transcriptional response to in vivo BCG-induced trained immunity. Cell Rep 42, 112487. 10.1016/j.celrep.2023.112487.

14. Stražar, M., Mourits, V.P., Koeken, V.A.C.M., de Bree, L.C.J., Moorlag, S.J.C.F.M., Joosten, L.A.B., van Crevel, R., Vlamakis, H., Netea, M.G., and Xavier, R.J. (2021). The influence of the gut microbiome on BCG-induced trained immunity. Genome Biol 22, 275. 10.1186/s13059-021-02482-0.

15. Pavan Kumar, N., Padmapriyadarsini, C., Rajamanickam, A., Marinaik, S.B., Nancy, A., Padmanaban, S., Akbar, N., Murhekar, M., and Babu, S. (2021). Effect of BCG vaccination on proinflammatory responses in elderly individuals. Sci Adv 7, eabg7181. 10.1126/sciadv.abg7181.

16. Fanucchi, S., Domínguez-Andrés, J., Joosten, L.A.B., Netea, M.G., and Mhlanga, M.M. (2021). The Intersection of Epigenetics and Metabolism in Trained Immunity. Immunity 54, 32–43. 10.1016/j.immuni.2020.10.011.

17. Pacis, A., Tailleux, L., Morin, A.M., Lambourne, J., MacIsaac, J.L., Yotova, V., Dumaine, A., Danckaert, A., Luca, F., Grenier, J.-C., et al. (2015). Bacterial infection remodels the DNA methylation landscape of human dendritic cells. Genome Res 25, 1801–1811. 10.1101/gr.192005.115.

18. Gupta, M.K., Peng, H., Li, Y., and Xu, C.-J. (2023). The role of DNA methylation in personalized medicine for immune-related diseases. Pharmacol Ther 250, 108508. 10.1016/j.pharmthera.2023.108508.

19. Pacis, A., Mailhot-Léonard, F., Tailleux, L., Randolph, H.E., Yotova, V., Dumaine, A., Grenier, J.-C., and Barreiro, L.B. (2019). Gene activation precedes DNA demethylation in response to infection in human dendritic cells. Proc. Natl. Acad. Sci. U.S.A. 116, 6938–6943. 10.1073/pnas.1814700116.

20. Bannister, S., Kim, B., Domínguez-Andrés, J., Kilic, G., Ansell, B.R.E., Neeland, M.R., Moorlag, S.J.C.F.M., Matzaraki, V., Vlahos, A., Shepherd, R., et al. (2022). Neonatal BCG vaccination is associated with a long-term DNA methylation signature in circulating monocytes. Sci Adv 8, eabn4002. 10.1126/sciadv.abn4002.

21. Takahashi, H., Kühtreiber, W.M., Keefe, R.C., Lee, A.H., Aristarkhova, A., Dias, H.F., Ng, N., Nelson, K.J., Bien, S., Scheffey, D., et al. (2022). BCG vaccinations drive epigenetic changes to the human T cell receptor: Restored expression in type 1 diabetes. Sci Adv 8, eabq7240. 10.1126/sciadv.abq7240.

22. Verma, D., Parasa, V.R., Raffetseder, J., Martis, M., Mehta, R.B., Netea, M., and Lerm, M. (2017). Anti-mycobacterial activity correlates with altered DNA methylation pattern in immune cells from BCG-vaccinated subjects. Sci Rep 7, 12305. 10.1038/s41598-017-12110-2.

23. Debisarun, P.A., Kilic, G., de Bree, L.C.J., Pennings, L.J., van Ingen, J., Benn, C.S., Aaby, P., Dijkstra, H., Lemmers, H., Domínguez-Andrés, J., et al. (2023). The impact of BCG dose and revaccination on trained immunity. Clin Immunol 246, 109208. 10.1016/j.clim.2022.109208.

24. Houseman, E., Accomando, W.P., Koestler, D.C., Christensen, B.C., Marsit, C.J., Nelson, H.H., Wiencke, J.K., and Kelsey, K.T. (2012). DNA methylation arrays as surrogate measures of cell mixture distribution. BMC Bioinformatics 13, 86. 10.1186/1471-2105-13-86.

25. Li, Y.-L., Zhao, H., and Ren, X.-B. (2016). Relationship of VEGF/VEGFR with immune and cancer cells: staggering or forward? Cancer Biol Med 13, 206–214. 10.20892/j.issn.2095-3941.2015.0070.

26. Chen, M.L., Ge, Z., Fox, J.G., and Schauer, D.B. (2006). Disruption of tight junctions and induction of proinflammatory cytokine responses in colonic epithelial cells by Campylobacter jejuni. Infect Immun 74, 6581–6589. 10.1128/IAI.00958-06.

27. Breeze, C.E., Reynolds, A.P., van Dongen, J., Dunham, I., Lazar, J., Neph, S., Vierstra, J., Bourque, G., Teschendorff, A.E., Stamatoyannopoulos, J.A., et al. (2019). eFORGE v2.0: updated analysis of cell type-specific signal in epigenomic data. Bioinformatics 35, 4767–4769. 10.1093/bioinformatics/btz456.

28. Hernando-Herraez, I., Evano, B., Stubbs, T., Commere, P.-H., Jan Bonder, M., Clark, S., Andrews, S., Tajbakhsh, S., and Reik, W. (2019). Ageing affects DNA methylation drift and transcriptional cell-to-cell variability in mouse muscle stem cells. Nat Commun 10, 4361. 10.1038/s41467-019-12293-4.

29. Choi, S.W., Mak, T.S.-H., and O’Reilly, P.F. (2020). Tutorial: a guide to performing polygenic risk score analyses. Nat Protoc 15, 2759–2772. 10.1038/s41596-020-0353-1.

30. Huang, H., Xiong, Q., Wang, N., Chen, R., Ren, H., Siwko, S., Han, H., Liu, M., Qian, M., and Du, B. (2018). Kisspeptin/GPR54 signaling restricts antiviral innate immune response through regulating calcineurin phosphatase activity. Sci Adv 4, eaas9784. 10.1126/sciadv.aas9784.

31. Wang, Z., Wu, J., Jiang, J., Ma, Q., Song, M., Xu, T., Liu, Y., Chen, Z., Bao, Y., Huang, M., et al. (2022). KIF2A decreases IL-33 production and attenuates allergic asthmatic inflammation. Allergy Asthma Clin Immunol 18, 55. 10.1186/s13223-022-00697-9.

32. de Vries, S., Benes, V., Naarmann-de Vries, I.S., Rücklé, C., Zarnack, K., Marx, G., Ostareck, D.H., and Ostareck-Lederer, A. (2021). P23 Acts as Functional RBP in the Macrophage Inflammation Response. Front Mol Biosci 8, 625608. 10.3389/fmolb.2021.625608.

33. Cheng, S.-C., Quintin, J., Cramer, R.A., Shepardson, K.M., Saeed, S., Kumar, V., Giamarellos-Bourboulis, E.J., Martens, J.H.A., Rao, N.A., Aghajanirefah, A., et al. (2014). mTOR- and HIF-1α-mediated aerobic glycolysis as metabolic basis for trained immunity. Science 345, 1250684. 10.1126/science.1250684.

34. Setoguchi, R., Matsui, Y., and Mouri, K. (2015). mTOR signaling promotes a robust and continuous production of IFN-γ by human memory CD8+ T cells and their proliferation. Eur J Immunol 45, 893–902. 10.1002/eji.201445086.

35. Tingley, D., Yamamoto, T., Hirose, K., Keele, L., and Imai, K. (2014). Mediation: R package for causal mediation analysis.

36. Wang, C., and Ruan, J. (2023). An ancient defense mechanism: Conservation of gasdermin-mediated pyroptosis. PLoS Biol 21, e3002103. 10.1371/journal.pbio.3002103.

37. Greenwood, C.S., Wynosky-Dolfi, M.A., Beal, A.M., and Booty, L.M. (2023). Gasdermins assemble; recent developments in bacteriology and pharmacology. Front Immunol 14, 1173519. 10.3389/fimmu.2023.1173519.

38. Cruz-Rangel, S., Melo, Z., Vázquez, N., Meade, P., Bobadilla, N.A., Pasantes-Morales, H., Gamba, G., and Mercado, A. (2011). Similar effects of all WNK3 variants on SLC12 cotransporters. Am J Physiol Cell Physiol 301, C601–608. 10.1152/ajpcell.00070.2011.

39. Lam, B.Y.H., Williamson, A., Finer, S., Day, F.R., Tadross, J.A., Gonçalves Soares, A., Wade, K., Sweeney, P., Bedenbaugh, M.N., Porter, D.T., et al. (2021). MC3R links nutritional state to childhood growth and the timing of puberty. Nature 599, 436–441. 10.1038/s41586-021-04088-9.

40. Gumus, E. (2018). Case report of four siblings in southeast Turkey with a novel RAB3GAP2 splice site mutation: Warburg micro syndrome or Martsolf syndrome? Ophthalmic Genet 39, 391–395. 10.1080/13816810.2018.1432065.

41. Quintin, J., Saeed, S., Martens, J.H.A., Giamarellos-Bourboulis, E.J., Ifrim, D.C., Logie, C., Jacobs, L., Jansen, T., Kullberg, B.-J., Wijmenga, C., et al. (2012). Candida albicans infection affords protection against reinfection via functional reprogramming of monocytes. Cell Host Microbe 12, 223–232. 10.1016/j.chom.2012.06.006.

42. Saeed, S., Quintin, J., Kerstens, H.H.D., Rao, N.A., Aghajanirefah, A., Matarese, F., Cheng, S.-C., Ratter, J., Berentsen, K., van der Ent, M.A., et al. (2014). Epigenetic programming of monocyte-to-macrophage differentiation and trained innate immunity. Science 345, 1251086. 10.1126/science.1251086.

43. Netea, M.G., Domínguez-Andrés, J., Barreiro, L.B., Chavakis, T., Divangahi, M., Fuchs, E., Joosten, L.A.B., van der Meer, J.W.M., Mhlanga, M.M., Mulder, W.J.M., et al. (2020). Defining trained immunity and its role in health and disease. Nat Rev Immunol 20, 375–388. 10.1038/s41577-020-0285-6.

44. Unnikrishnan, A., Freeman, W.M., Jackson, J., Wren, J.D., Porter, H., and Richardson, A. (2019). The role of DNA methylation in epigenetics of aging. Pharmacol Ther 195, 172–185. 10.1016/j.pharmthera.2018.11.001.

45. Das, J., Verma, D., Gustafsson, M., and Lerm, M. (2019). Identification of DNA methylation patterns predisposing for an efficient response to BCG vaccination in healthy BCG-naïve subjects. Epigenetics 14, 589–601. 10.1080/15592294.2019.1603963.

46. Kleinnijenhuis, J., Quintin, J., Preijers, F., Benn, C.S., Joosten, L.A.B., Jacobs, C., van Loenhout, J., Xavier, R.J., Aaby, P., van der Meer, J.W.M., et al. (2014). Long-lasting effects of BCG vaccination on both heterologous Th1/Th17 responses and innate trained immunity. J Innate Immun 6, 152–158. 10.1159/000355628.

47. Wang, D., Wu, Z., Zhao, C., Yang, X., Wei, H., Liu, M., Zhao, J., Qian, M., Li, Z., and Xiao, J. (2021). KP-10/Gpr54 attenuates rheumatic arthritis through inactivating NF-κB and MAPK signaling in macrophages. Pharmacol Res 171, 105496. 10.1016/j.phrs.2021.105496.

48. Watanabe, T., and Sato, K. (2020). Roles of the kisspeptin/GPR54 system in pathomechanisms of atherosclerosis. Nutr Metab Cardiovasc Dis 30, 889–895. 10.1016/j.numecd.2020.02.017.

49. Murphy, D.M., Mills, K.H.G., and Basdeo, S.A. (2021). The Effects of Trained Innate Immunity on T Cell Responses; Clinical Implications and Knowledge Gaps for Future Research. Front Immunol 12, 706583. 10.3389/fimmu.2021.706583.

50. Stensballe, L.G., Nante, E., Jensen, I.P., Kofoed, P.-E., Poulsen, A., Jensen, H., Newport, M., Marchant, A., and Aaby, P. (2005). Acute lower respiratory tract infections and respiratory syncytial virus in infants in Guinea-Bissau: a beneficial effect of BCG vaccination for girls community based case-control study. Vaccine 23, 1251–1257. 10.1016/j.vaccine.2004.09.006.

51. Biering-Sørensen, S., Jensen, K.J., Monterio, I., Ravn, H., Aaby, P., and Benn, C.S. (2018). Rapid Protective Effects of Early BCG on Neonatal Mortality Among Low Birth Weight Boys: Observations From Randomized Trials. J Infect Dis 217, 759–766. 10.1093/infdis/jix612.

52. Zakharova, L., Sharova, V., and Izvolskaia, M. (2020). Mechanisms of Reciprocal Regulation of Gonadotropin-Releasing Hormone (GnRH)-Producing and Immune Systems: The Role of GnRH, Cytokines and Their Receptors in Early Ontogenesis in Normal and Pathological Conditions. Int J Mol Sci 22, 114. 10.3390/ijms22010114.

53. Yang, J., Qi, F., Gu, H., Zou, J., Yang, Y., Yuan, Q., and Yao, Z. (2016). Neonatal BCG vaccination of mice improves neurogenesis and behavior in early life. Brain Res Bull 120, 25–33. 10.1016/j.brainresbull.2015.10.012.

54. Nag, K., and Chaudhary, A. (2009). Mediators of Tyrosine Phosphorylation in Innate Immunity: From Host Defense to Inflammation onto Oncogenesis. Curr Signal Transduct Ther 4, 76–81. 10.2174/15743620978816750.

55. Huda, M.N., Ahmad, S.M., Alam, M.J., Khanam, A., Afsar, M.N.A., Wagatsuma, Y., Raqib, R., Stephensen, C.B., and Laugero, K.D. (2019). Infant cortisol stress-response is associated with thymic function and vaccine response. Stress 22, 36–43. 10.1080/10253890.2018.1484445.

56. Aryee, M.J., Jaffe, A.E., Corrada-Bravo, H., Ladd-Acosta, C., Feinberg, A.P., Hansen, K.D., and Irizarry, R.A. (2014). Minfi: a flexible and comprehensive Bioconductor package for the analysis of Infinium DNA methylation microarrays. Bioinformatics 30, 1363–1369. 10.1093/bioinformatics/btu049.

57. Pidsley, R., Zotenko, E., Peters, T.J., Lawrence, M.G., Risbridger, G.P., Molloy, P., Van Djik, S., Muhlhausler, B., Stirzaker, C., and Clark, S.J. (2016). Critical evaluation of the Illumina MethylationEPIC BeadChip microarray for whole-genome DNA methylation profiling. Genome Biol 17, 208. 10.1186/s13059-016-1066-1.

58. Zhou, W., Laird, P.W., and Shen, H. (2017). Comprehensive characterization, annotation and innovative use of Infinium DNA methylation BeadChip probes. Nucleic Acids Res 45, e22. 10.1093/nar/gkw967.

59. Touleimat, N., and Tost, J. (2012). Complete pipeline for Infinium(®) Human Methylation 450K BeadChip data processing using subset quantile normalization for accurate DNA methylation estimation. Epigenomics 4, 325–341. 10.2217/epi.12.21.

60. McLean, C.Y., Bristor, D., Hiller, M., Clarke, S.L., Schaar, B.T., Lowe, C.B., Wenger, A.M., and Bejerano, G. (2010). GREAT improves functional interpretation of cis-regulatory regions. Nature Biotechnology 28, 495–501. 10.1038/nbt.1630.

61. Shabalin, A.A. (2012). Matrix eQTL: ultra fast eQTL analysis via large matrix operations. Bioinformatics 28, 1353–1358. 10.1093/bioinformatics/bts163.

