## Supplementary material for "Epigenetic Reprogramming Mediates Monocyte and Heterologous T Cell-derived Cytokine Responses after BCG Vaccination": Figure S

### Supplemental Figures

**Figure S1. Heatmap of association between covariates, estimated cell counts and DNA methylation at T14 (A) and T90 (B)**

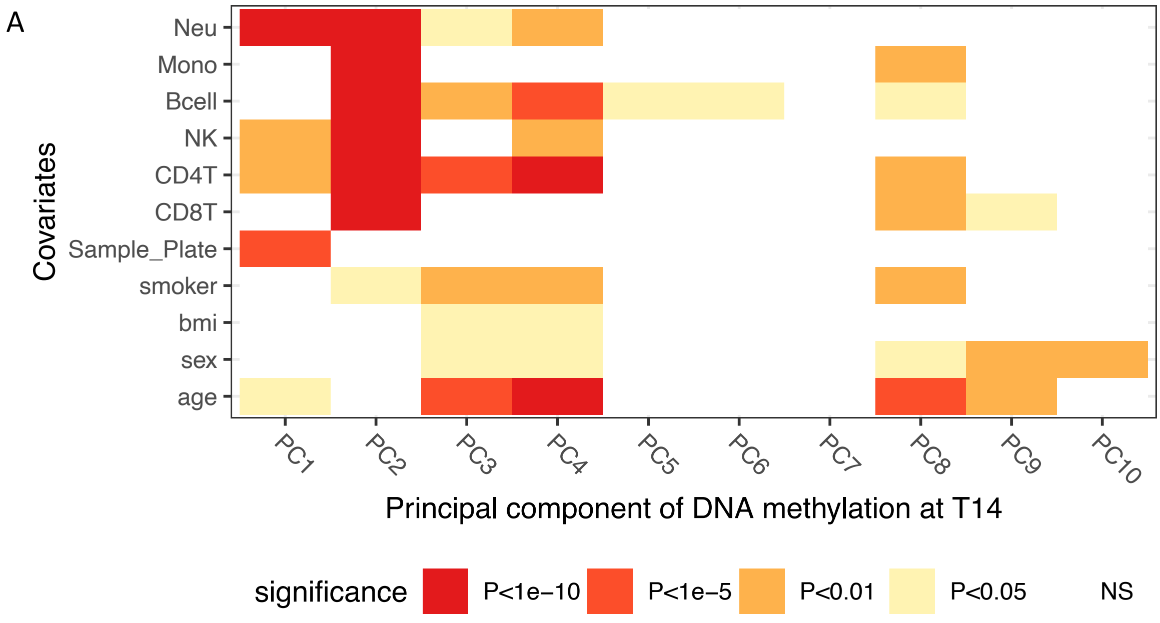


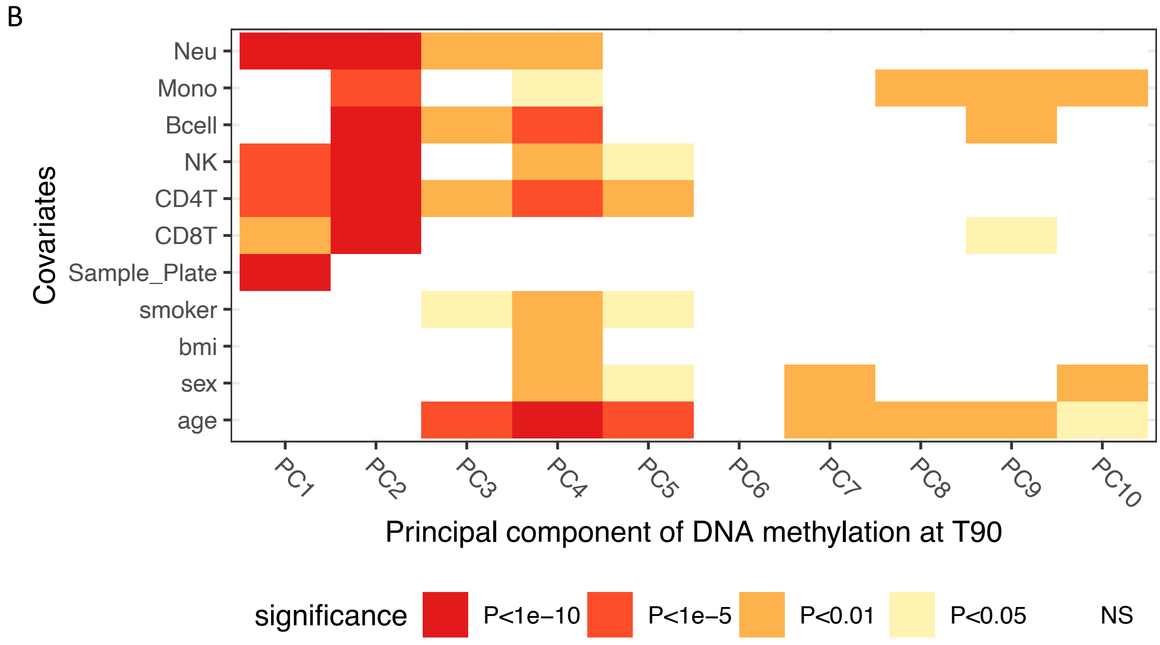

**Figure S2. BCG-induced DNA methylation changes.** (A)Dot plot showing the pathway enrichment of genes annotated to the identified CpG sites (FDR<0.05). The color indicates the source dataset used in the enrichment analysis, and the size of dot shows the number of genes enriched to the pathway. Heatmap of correlations between identified 15 CpG sites and estimated cell proportions (B), as well as cell counts of neutrophil sub cell types measured by flow cytometry (C), at T0, T14 and T90 (from top to bottom) respectively. Cell colors indicate the correlation coefficient of spearman correlation, and the asterisks indicate the significance of the correlation (* P<0.05, ** P<0.01, ***P<0.001).

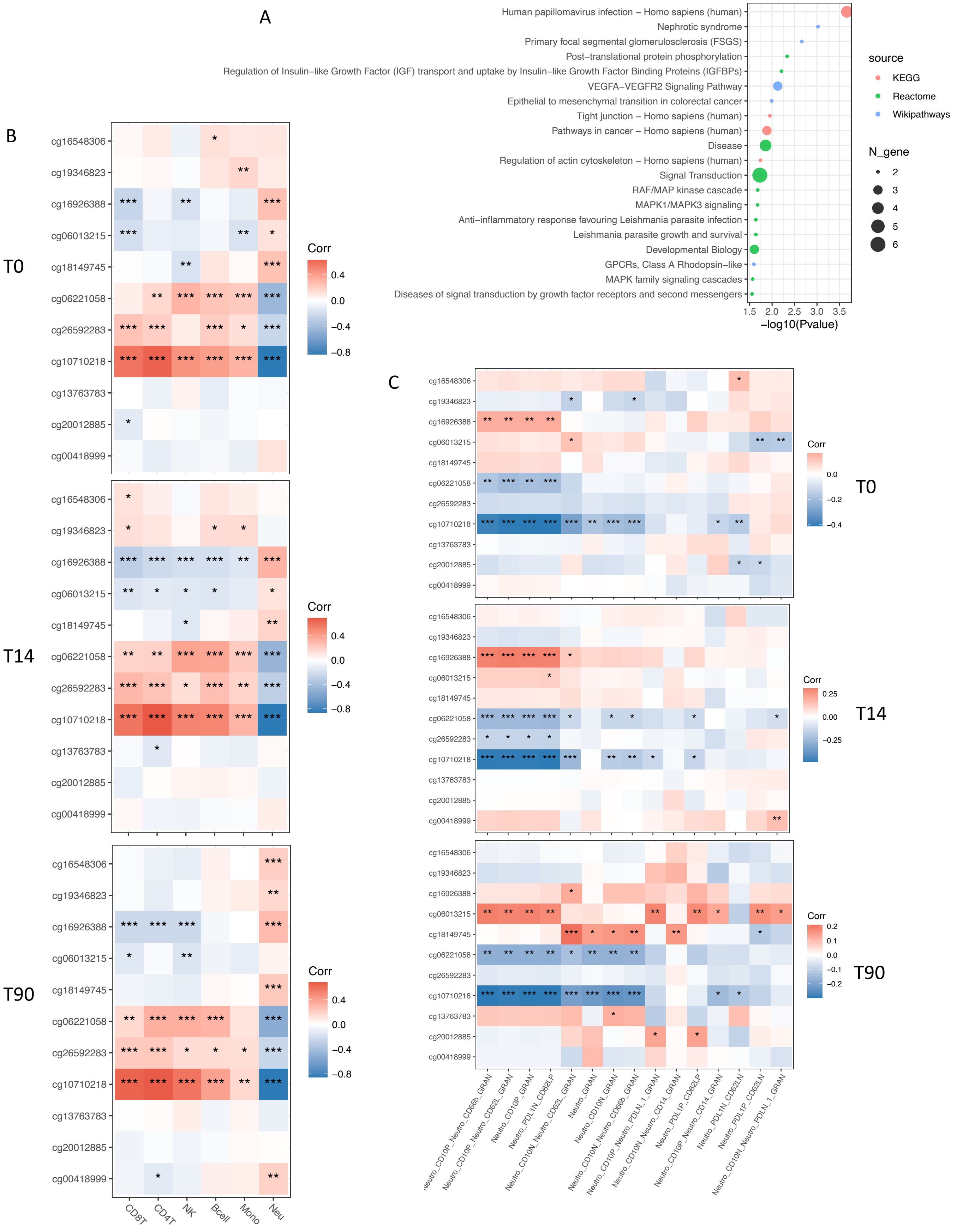

**Figure S3. Short- and Long-term DNA methylation changes induced by BCG vaccination.** Heatmap of hierarchical clustering on DNA methylation changes of identified CpG sites at T14-T0 (A), T90-T0 (B) and T90-T14 (C), which are represented as the differences in DNA methylation beta values. t21: beta value differences between T14 and T0, t32: differences between T90 and T14, t31:differences between T90 and T0.

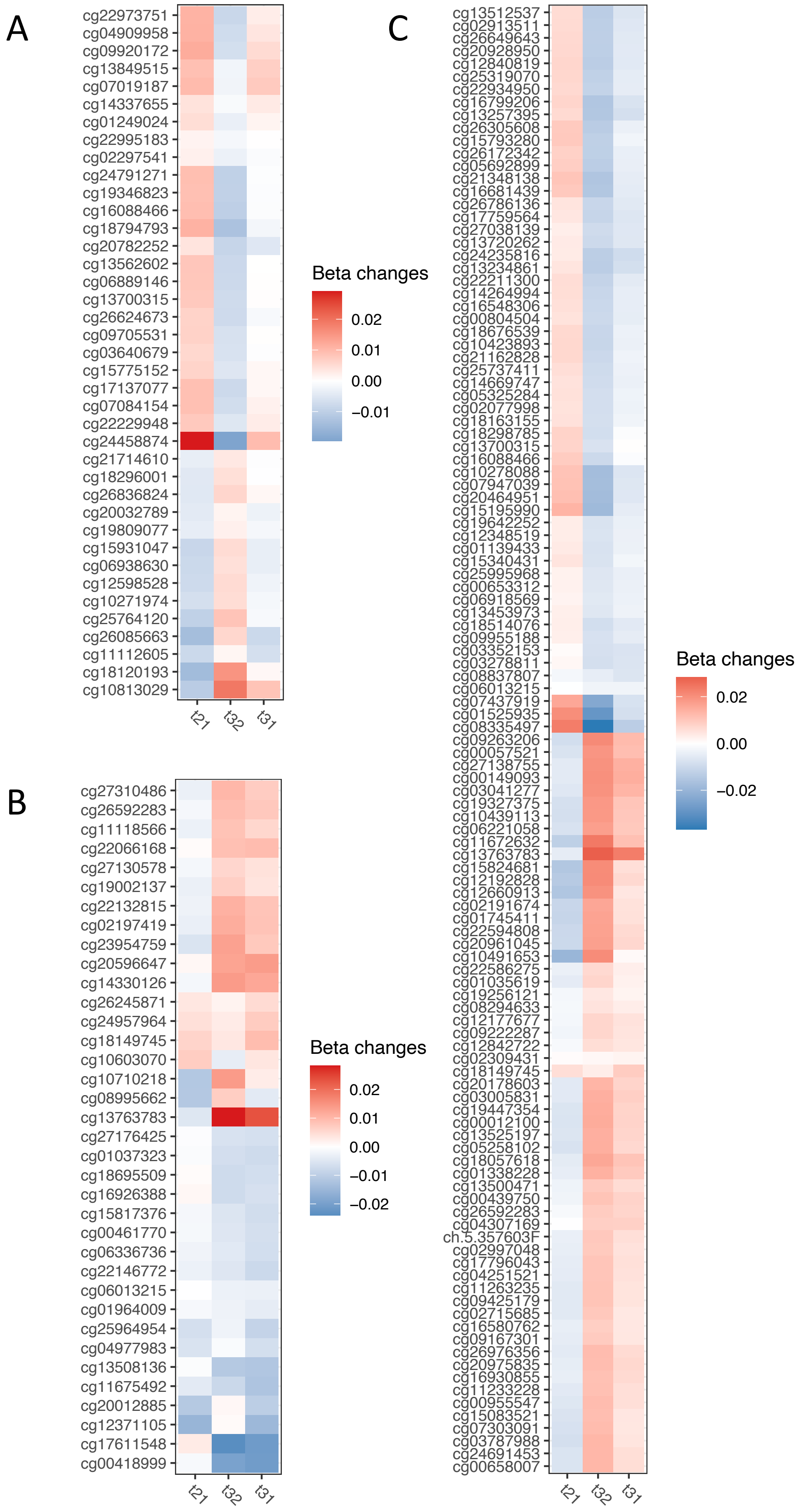

**Figure S4. Short- and Long-term DNA methylation changes induced by BCG vaccination.** (A-C) Boxplot plots shows the average changes of hyper and hypo methylated CpG sites identified from model T14-T0 (A, short-term effect), T90-T0 (B, long-term effect), and T90-T14 (C, late effect). Paired t-test was used to assess the significant differences of average methylation changes over time points. (D-E) Dot plot showing the pathway enrichment of genes annotated to the identified CpG sites (P<1e-5) from the model T14-T0 (D), T90-T0(E) and T90-T14 (F).

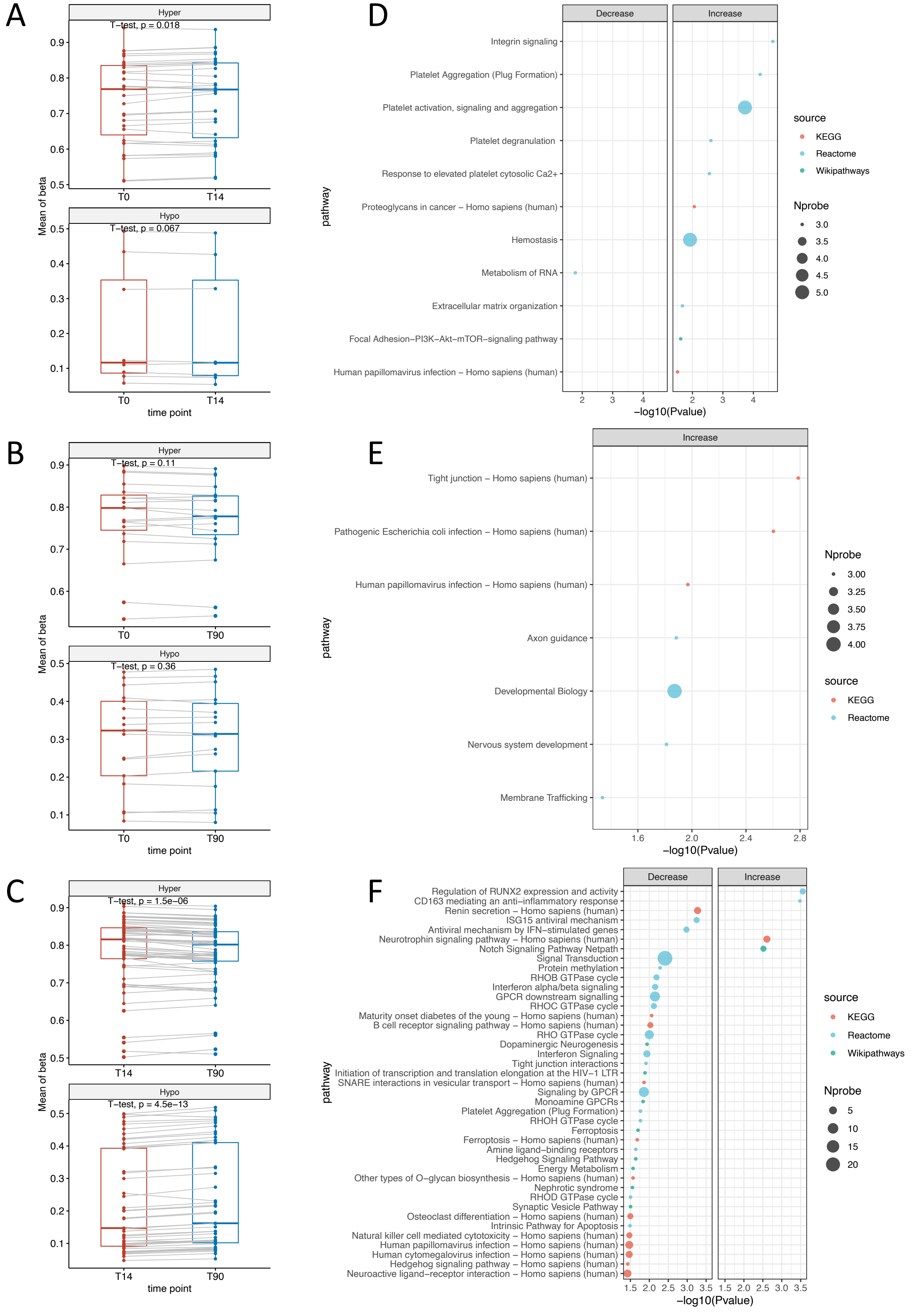

**Figure S5. BCG-induced methylation changes and inflammatory protein changes.** (A) Heatmap of hierarchical clustering on Spearman correlation patterns between DNA methylation changes and protein changes at T14 compared with T0 (A) and T90 compared with T0 (B). CpG sites were identified by comparing DNA methylation level at T14 to T0 (A) and T90 to T0 (B). Group of decrease and increase indicates the direction of DNA methylation levels at T14 compared with T0 (A) and T90 compared with T0 (B). Proteins were reported in the previous study that showed significant decrease after BCG vaccination. Differences of methylation level and fold changes of proteins at these two time points mentioned were used in the analyses. CpG-protein pairs with P value<0.05 was marked by #, and the -log10 Pvalue * sign of correlation coefficient was plotted in each cell.

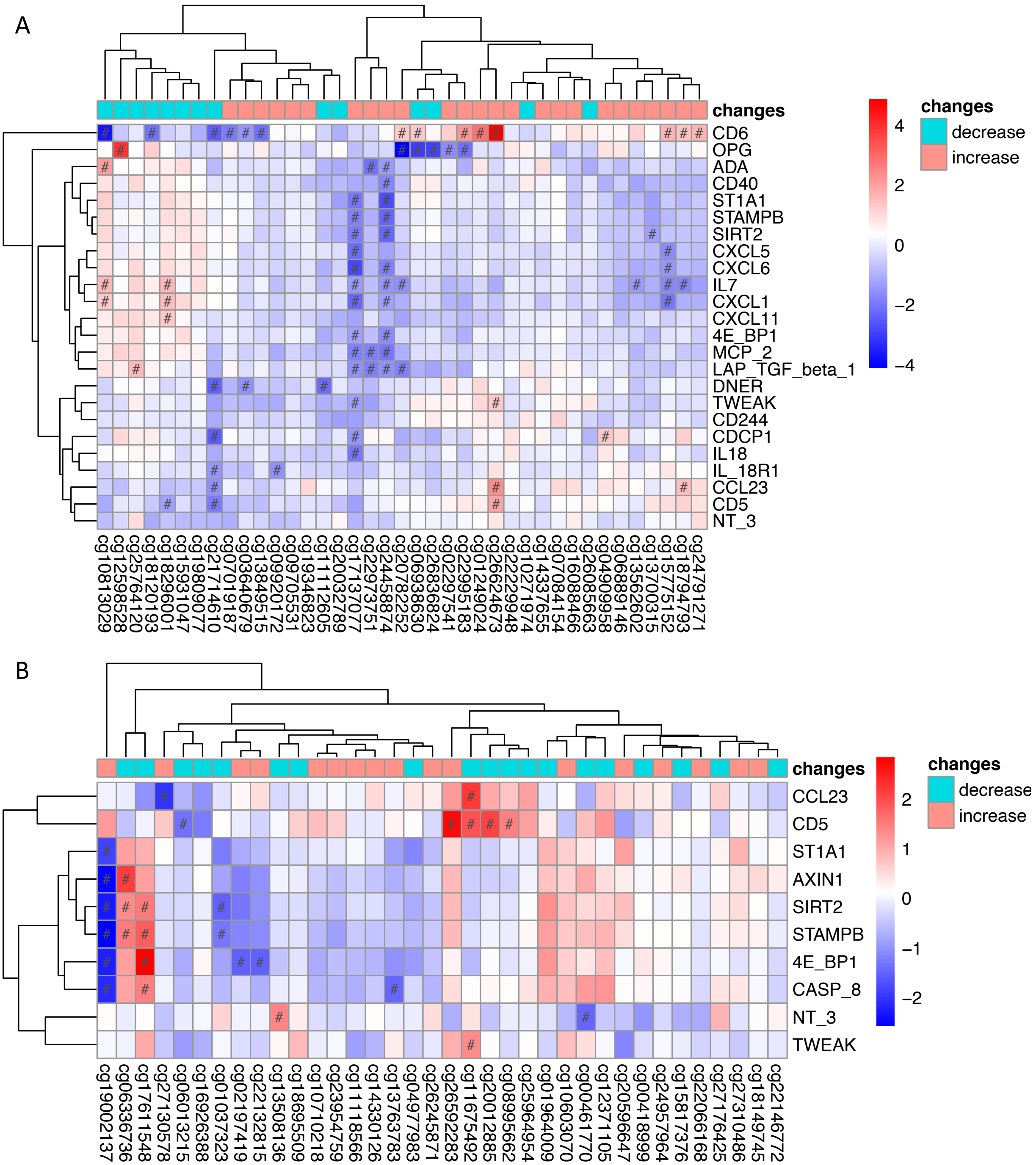

**Figure S6. Annotation of CpG sites associated with ex-vivo IFN-γ production changes upon *S. aureus* stimulation** (A) Dot plot showing the pathway enrichment of genes annotated to the identified CpG sites (FDR<0.05). (B) Bar plot showing the number of CpG sites associated with other traits that were available in the EWAS catalog (http://www.ewascatalog.org ). (C) Variance of *ex-vivo* IFN-γ response explained by randomly selected CpG sites and metabolites for 1000 times. The red dashed line represent the variance explained by CpG sites or metabolites that significantly correlated with *ex-vivo* IFN-γ response (same as the value in Figure 3B). (D) The enrichment of SNPs around the identified CpG sites to cytokine QTL. Cytokine QTL p-values of those SNPs located 250 kbp upstream or downstream of those CpG sites were extracted, and plotted against random cytokine QTL p-values.

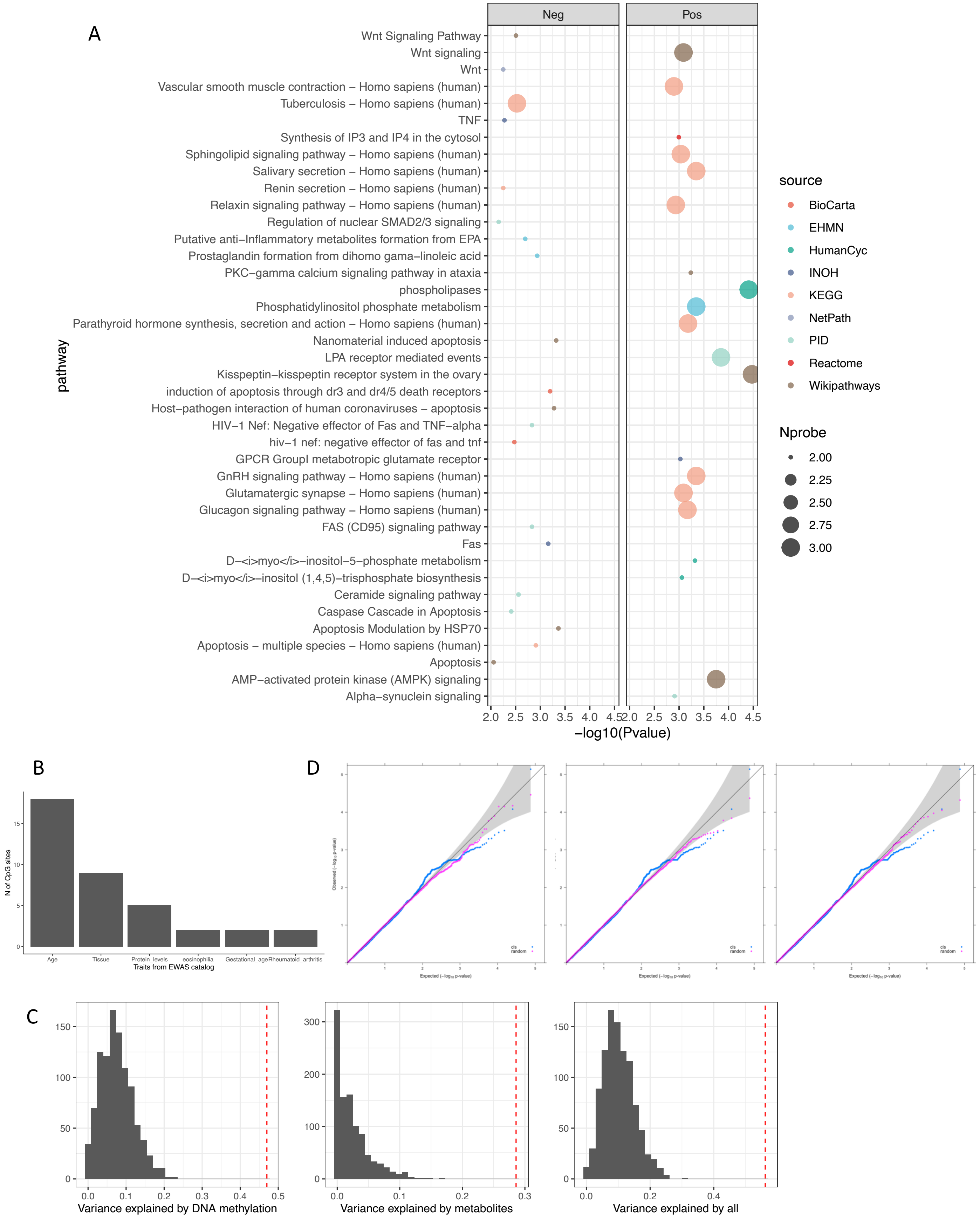

**Figure S7. Graphical summary of association analyses applied in this study.** (A) Summary of three main analyses in this study: Analysis1, BCG induced DNA methylation changes, inducing short-term changes (model T14-T0), long-term changes (model T90-T0) and late-effect (T90-T14); Analysis2, Baseline DNA methylation associated with trained immunity (TI, *ex-vivo* cytokine production changes upon *S. aureus* stimulation before and after BCG vaccination); Analysis2, the association between DNA methylation changes TI. Numbers of specific significance threshold are shown for each analysis. (B) Numbers of significant CpG sites of Analysis1 (P<1e-5) that passed P<0.05 in Analysis2. (C) Numbers of significant CpG sites of Analysis1 (P<1e-5) that passed P<0.05 in Analysis3.

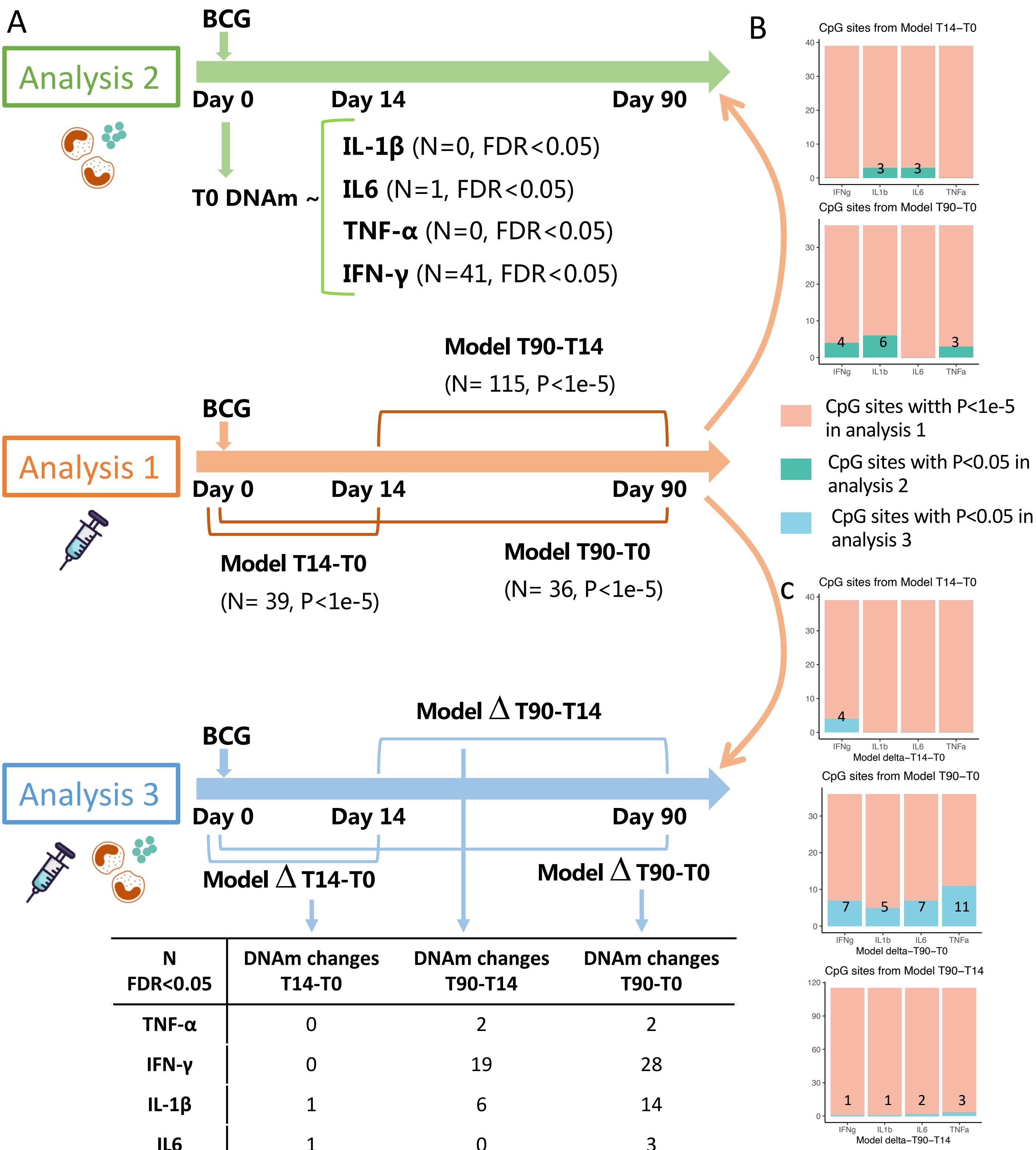


| N FDR<0.05 | DNAm changes T14-T0 | DNAm changes T90-T14 | DNAm changes T90-T0 |
|---|---|---|---|
| TNF-α | 0 | 2 | 2 |
| IFN-γ | 0 | 19 | 28 |
| IL-1β | 1 | 6 | 14 |
| IL6 | 1 | 0 | 3 |

**Figure S8.** Dot plot showing the pathway enrichment of genes annotated to CpG sites identified from the association analyses between the changes in DNA methylation after BCG vaccination and trained immunity (TI, *ex-vivo* cytokine production changes upon *S. aureus* stimulation before and after BCG vaccination) (P<1e-5, Analysis3, model ∆ T90-T0 ). (A-D) represents different cytokines respectively.

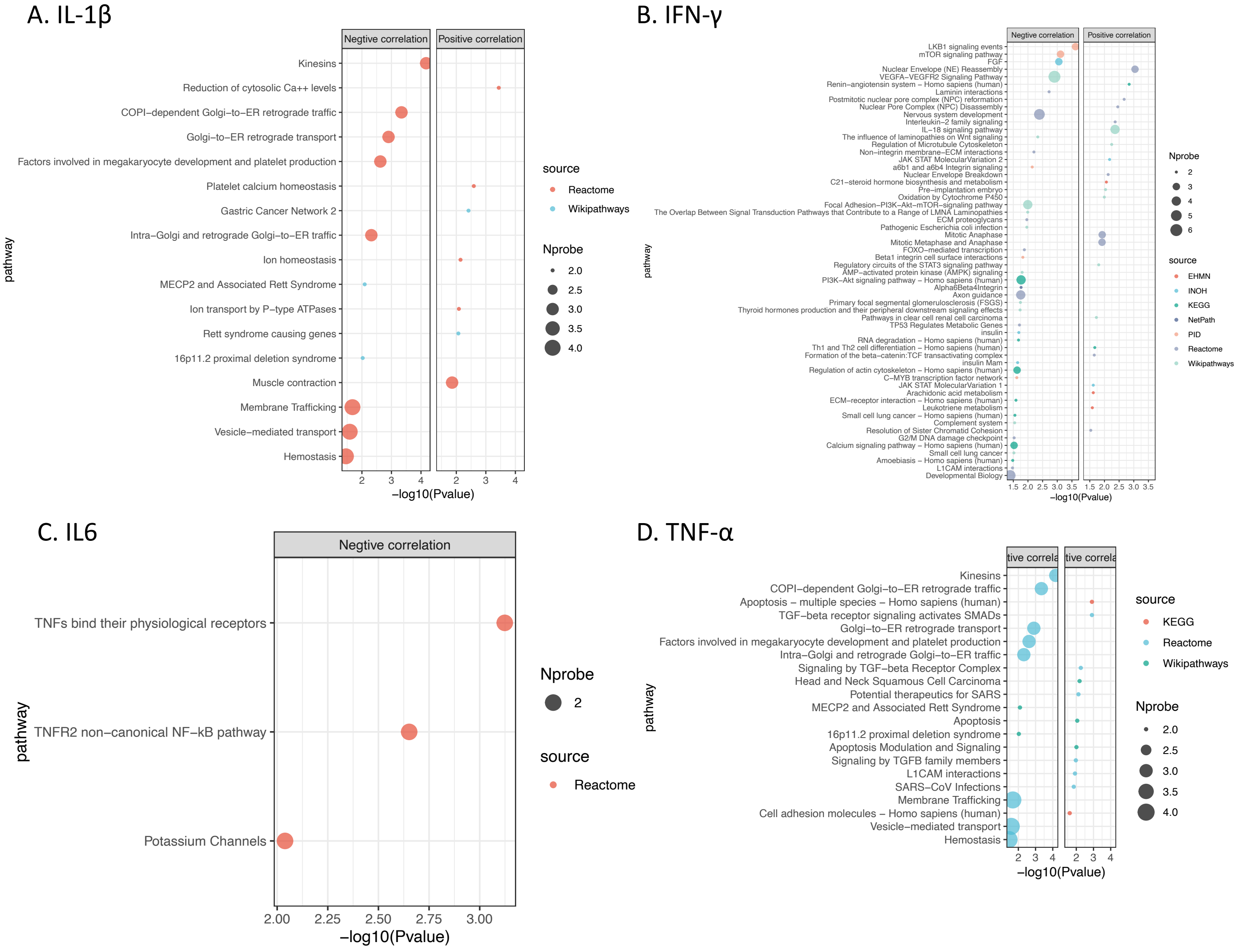

**Figure S9 Causal relationship inference by bidirectional mediation analysis.** (A) Sankey diagram showing the inferred causal relationship network of Direction2 (DNAm-C mediates the effect of SNP on TI) with mediation P value <0.05. (B-D) Example of causal relationships between SNP, DNAm-C and TI inferred by bidirectional mediation analysis. The beta coefficient and significance are labeled at each edge and the proportions of mediation effect are labeled at the center of ring charts. See also Table S14. DNAm-C: DNA methylation changes. TI: trained immunity.

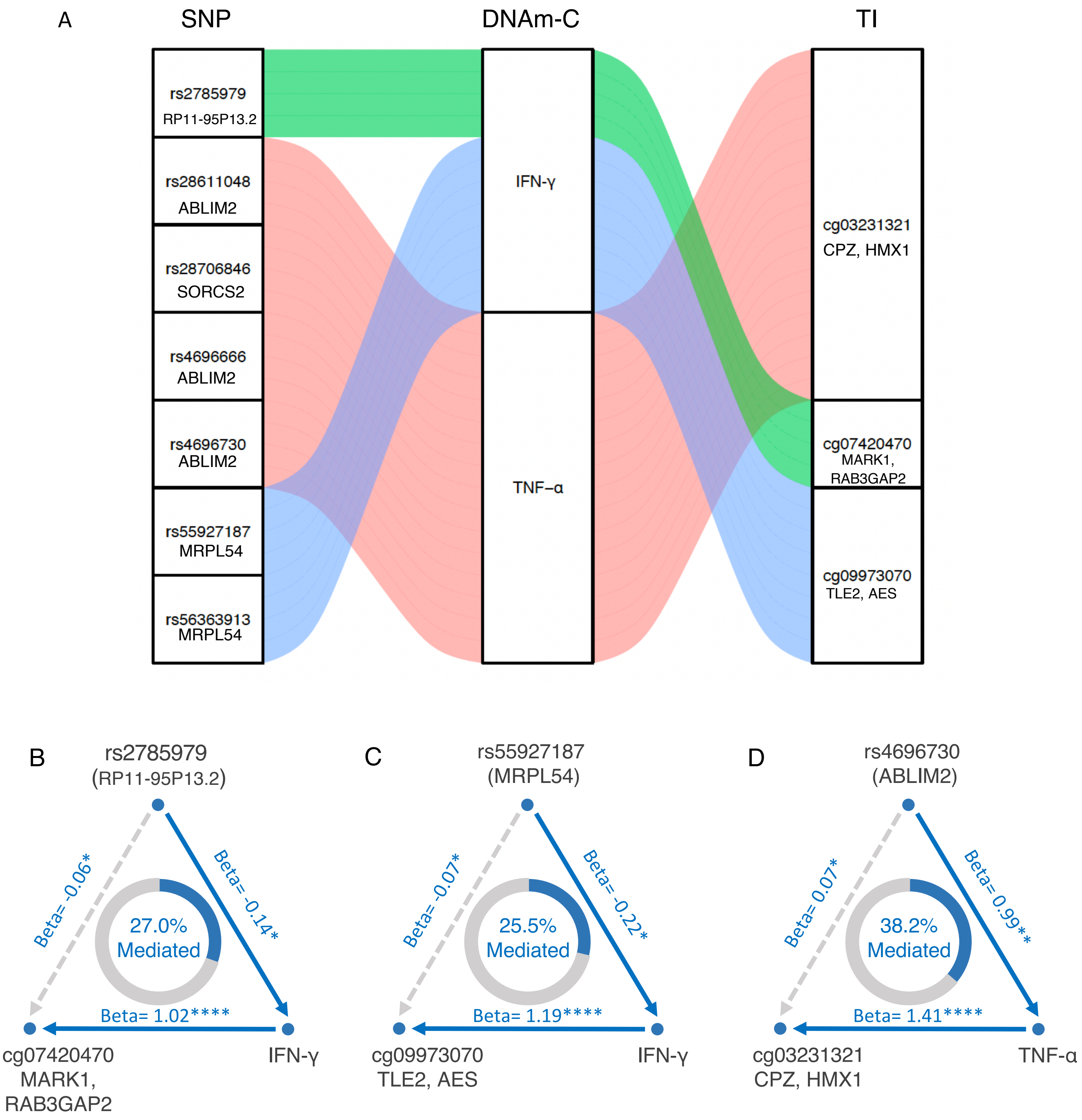

**Figure S10. Sex-specific effect on the DNA methylation changes induced by BCG vaccination.** Change patterns of CpG sites identified which assessed the DNA methylation changes at day14 and day0 (T14-T0, A), day90 and day0 (T90-T0, B) with P<1e-5 in male (upper panel) and female (lower panel) respectively. (C-D), Dot plot showing the pathway enrichment of genes annotated to the identified CpG sites from A-B.

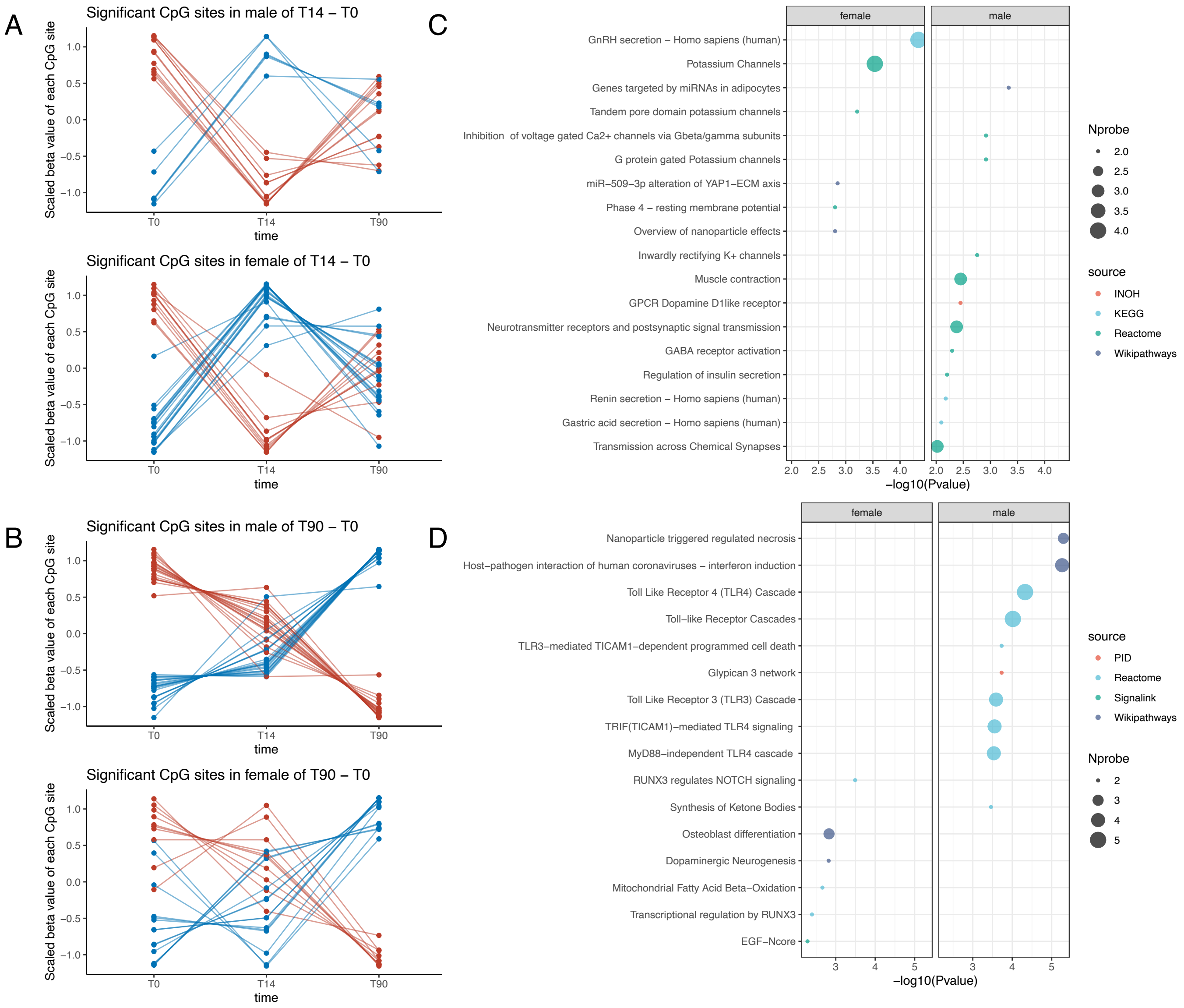

**Figure S11.** Dot plot showing the pathway enrichment of genes annotated to the CpG sites associated with *ex-vivo* IFN-γ production changes upon *S. aureus* stimulation before and after BCG vaccination in male and female.

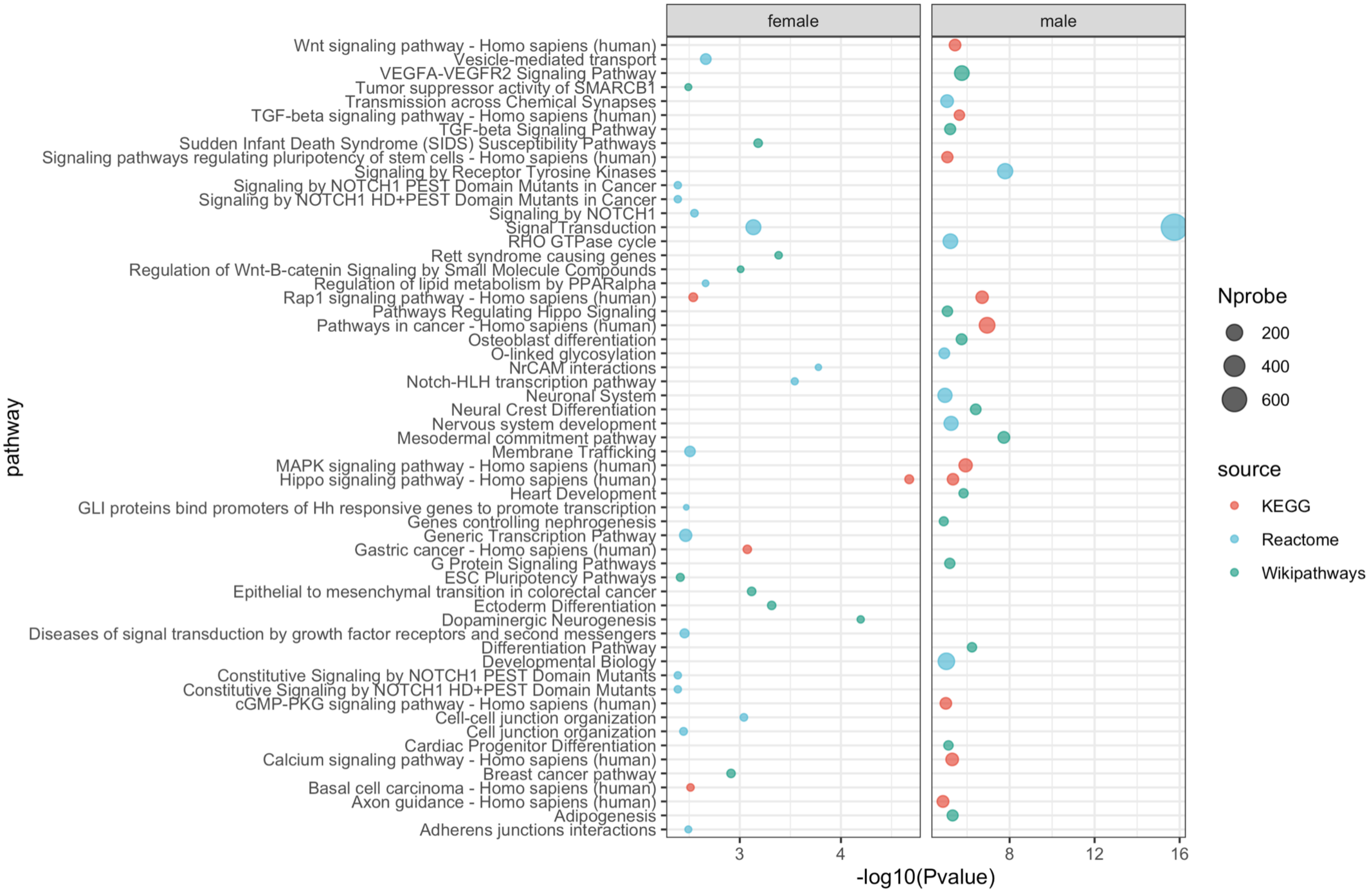
